# Longitudinal changes of health-related quality of life, digital media use, and physical activity in children between 4-17 years during the COVID-19 pandemic in Germany

**DOI:** 10.64898/2026.11.24.26368819

**Authors:** Claudia Lazarides, Janis Fiedler, Leon Klos, Carmen Volk, Claudia Niessner, Manuel C. Voelkle, Alexander Woll

## Abstract

**Background:** Evidence in youth on whether pandemic-related changes in physical activity (PA), recreational screen time (ST), and health-related quality of life (HRQoL) persisted after the COVID-19 pandemic is limited. In a longitudinal study, we examined changes from pre-to post-pandemic and whether these changes differed by age at pre-pandemic.

**Methods:** Longitudinal data were obtained from 945 participants (49% female) aged 4-17 years pre-pandemic in the German Motorik-Modul Study. HRQoL, ST, everyday PA, and the number of days per week with at least 60 minutes of PA (WHO PA days) were assessed pre-pandemic (2018–2020), during both German lockdowns (2020 and 2021), and post-pandemic after restrictions ended (2023). Covariate-adjusted random-intercept mixed-effects models tested pre-to post-pandemic changes and moderation by baseline age. Lockdown trajectories were explored using successive contrasts.

**Results:** Compared with pre-pandemic levels, post-pandemic HRQoL decreased (B = −5.92, 95% CI [−6.82, −5.03]), everyday PA decreased by 31.15 minutes per day (95% CI [−36.40, −25.91]), and WHO PA days decreased by 0.27 days per week (95% CI [−0.43, −0.11]). ST increased by 75.17 minutes per day (95% CI [66.65, 83.69]; all *p* ≤ .001). Age moderated changes in HRQoL, ST, and everyday PA, but not WHO PA days. Children aged 6-13 years showed the greatest ST increases. Children younger than 11 years showed the largest everyday PA declines. Additionally explored trajectories varied across lockdowns and were moderated by age prior to the pandemic.

**Conclusions:** Children’s and adolescents’ PA, ST, and HRQoL had not returned to pre-pandemic levels one year after COVID-19 restrictions were lifted. Developmentally tailored monitoring and interventions may be required, particularly for younger and school-aged children in face of ongoing global crises.

## Introduction

### Relevance

The importance of health-related aspects of children’s and adolescents’ life such as health-related quality of life (HrQoL) and health behaviours (recreational screen time, ST; physical activity; PA), and their effect on children’s health have been extensively studied. Impairments in HrQoL have been linked to increased mortality in adulthood (Kaplan et al., 2007). High amounts of ST have been under investigation to affect children’s health negatively. However, findings have been inconsistent (Valkenburg et al., 2022). The positive effect of PA on health has been well described in the literature (Aston, 2018; Bull et al., 2020). Impairments in these health-related aspects of children’s life (HrQoL, ST, PA) potentially have long lasting negative effects on health, setting the stage for negative health trajectories in later life by means of developmental programming (Bianco-Miotto et al., 2017; Suzuki, 2018). Therefore, longitudinal changes in HrQoL, ST, and PA in the face of adversity (Boluda-Verdú et al., 2022; Kästner et al., 2022) should be identified as they may function as early warning signs for youth at risk for health impairments in later life, such as mental health issues, chronic disease, and obesity.

Importantly, the exposure to stressors (e.g., global crisis during COVID-19 pandemic) might have differential effects on health-related aspects and an individual’s health. Considering sensitive periods of human development the impact of stress might be different depending on the individual’s developmental stage (Lupien et al., 2009; Weiss & Wagner, 1998). Potentially, younger children or children at transitions points (e.g. transitioning from primary school to secondary school, kindergarten to primary school, puberty) might be differentially affected and therefore display differing changes in HrQoL, ST, and PA in the face of adversity.

### COVID-19 Pandemic as historic event

As a historic event, the COVID-19 pandemic and the temporary policy changes including restrictions on movement in public spaces, contact to others, and closure of public and private institutions impacted children’s and adolescents’ everyday life (Berner-Rodoreda et al., 2023). Restrictions in Germany were placed upon citizens in the form of two major lockdowns (Piesch et al., 2024) where HrQoL was impacted by multiple different causes such as fear of the health-related consequences of the virus and uncertainty of the future (Pelletier et al., 2021). Education-related and leisure-time activities transitioned to online settings, and organized opportunities for PA were limited. Aside of negative effects of the pandemic on youth mental health (Wolf & Schmitz, 2024), researchers reported impaired HrQoL, increased ST, and reduced sport PA but increased everyday PA from pre-pandemic times to Lockdown 1 using data of a nationwide, representative cohort study on children’s PA, motor performance and health (Wunsch et al., 2021).

### Research Gap

While there are a number of studies that underscore these developments *during* the pandemic (Rossi et al., 2021) and found that compared to pre-pandemic, HrQoL decreased (Vogel et al., 2021; Watson et al., 2023), ST increased (Haug et al., 2022; Hedderson et al., 2023; Helgadóttir et al., 2023), and PA decreased (Helgadóttir et al., 2023; O’Kane et al., 2021; Watson et al., 2023). Longitudinal studies reporting post-pandemic levels of HrQoL, ST, and PA are scarce. Previous studies not only lack assessments of the time that could be defined as “post-pandemic” broadly, the cited body of research also only included convenience samples, only measurements across longer periods during the pandemic as well as assessments after lockdowns rather than during these specific periods of restrictions. Taken together, these study designs limit the ability to infer how children and adolescents were doing in the aftermath of the pandemic compared to the pre-pandemic period and during the pandemic with regard to HrQoL, ST, and PA.

### Current study

To address this issue, the longitudinal nationwide, population-based Motorik-Modul (MoMo) Study which monitors PA, motor performance, and health in a nationally representative cohort of children and adolescents since 2003 in several assessment waves (Schmidt et al. 2020; Woll et al., 2011, 2017) adapted its data-collection procedures during the COVID-19 pandemic. For this analysis, Wave 3 serves as the pre-pandemic baseline, representing the last in-person assessment before the onset of COVID-19. Following Wave 3, three additional online assessments were conducted to capture the impact of the pandemic. The first two online assessments coincided with Germany’s nationwide COVID-19 lockdowns: Online Assessment 1 during the first lockdown (mid-March to early May 2020), and Online Assessment 2 during the second lockdown (November 2020 through spring 2021). A third online assessment was administered approximately two years after the end of the second lockdown (2022–2023), providing a post-pandemic measurement point. This longitudinal dataset allows us to examine changes in HrQoL, ST, and PA from the pre-pandemic to the post-pandemic period.

In this study, we examined the changes in HrQoL, ST and indicators of PA in a population-based sample of children and adolescents between (4-17y) living in Germany from 2018 until 2023. We investigated whether these changes varied by developmental stage, operationalized as age before the pandemic. We tested two hypotheses. First (H1), we hypothesized that, compared with pre-pandemic levels, HrQoL, everyday PA, and WHO PA days would be significantly lower and ST significantly higher post-pandemic, after adjusting for covariates previously associated with these outcomes independently of the passage of time (Thorpe & Gutman, 2023). Second (H2), we hypothesized that pre-pandemic age would moderate changes in HrQoL, ST, everyday PA, and WHO PA days over time. Finally, in an exploratory analysis, we examined successive differences in these outcomes across the four assessment points: pre-pandemic, lockdown1, lockdown2, and post-pandemic.

## Methods

### 1.1 Study design and participant characteristics

The MoMo Study is an epidemiological sub-study of the German Health Interview and Examination Survey for Children and Adolescents (KiGGS) (Mauz et al., 2020) and focuses on the on-site/in-person assessment of motor performance, PA, and health in children and adolescents. Additional information on the design and the sampling procedure of the MoMo Study is described elsewhere (Wagner et al., 2014; Woll et al., 2021). The MoMo Study was conducted in accordance with the Declaration of Helsinki. Ethics approval was obtained by the Charité Universitätsmedizin Berlin ethics committee, the University of Konstanz, and the ethics committee of the Karlsruhe Institute of Technology. The Federal Commissioner for Data Protection and Freedom of Information was informed about the study and approved it.

The current investigation uses data of wave 3 of the MoMo Study collected between 2018 until 2020 as the pre-pandemic baseline. Due to the governmental restrictions following the global COVID-19 pandemic in-person study visits including written responses to the health questionnaire were terminated in March 2020 preemptively. Following the governmental restrictions starting from 22^nd^ March 2020 onwards, all participants of wave 3 were invited to participate in the following online surveys collecting data on PA, ST, and HrQoL. The first online assessment took place during *Lockdown I* (governmental restriction 22^nd^ March – 6^th^ May, online assessment April 20, 2020 – May 1, 2020). The second online assessment was conducted during Lockdown II (02^nd^ November soft lockdown, 16^th^ December strict restrictions in place until early March 2021 with gradual release of restrictions, online assessment January 29, 2021 – February 14, 2021). The post-pandemic assessment took place with release of all restrictions in February 7-19, 2023 (end of all federally mandated mask requirements in everyday public life, except for health care institutions). The current investigation focuses in the comparison of wave 3 (pre-pandemic), to the post-pandemic follow-up and explores the trajectories through the lockdowns.

### 1.2 Measures

#### Outcomes

##### Health related quality of life

HrQoL was measured with the German version of the KIDSCREEN-10 (Ravens-Sieberer et al., 2014). The KIDSCREEN-10 is a short self-report measure assessing health related quality of life designed as a parent and child self-report. The scale showed sufficient psychometric reliability and validity in previous studies (Ravens-Sieberer et al., 2010). In the present study, reliability of the pre– and post-assessment was good (pre: Cronbach’s alpha = .75, post: Cronbach’s alpha =.80). Responses to 10 statements were given on a 5-points Likert-scale with two different response formats: a) from *not at all* to *extremely* (e.g., *Have you felt fit and well?*), or b) *never* to *always* (e.g., *Have your parent(s) treated you fairly?*). Items 3 (*Have you felt sad?*) and 4 (*Have you felt lonely?*) were recoded. Subsequently, sum scores of all responses to the 10 items were calculated for each assessment point. Sum scores were then transformed to standardized T-scores with norm data from German children and adolescents in 2019 to 2021 (Barkmann et al., 2021). Higher T-scores indicated higher HrQoL.

##### Recreational screen time

ST was measured through a questionnaire evaluating the amount of time spent watching television (TV), playing games on any device (Gaming), and using the internet for recreational use (Internet). The questions differentiated between weekdays and weekends using an 8-point scale including (almost) never, 15 min per day, 30 min per day, 1 h per day, 2 h per day, 3 h per day, and 4 h per day (Mathers et al., 2009). An index of total recreational screen time was calculated summarizing weekdays and weekend days at a 5:2 ratio.

##### Physical Activity

The assessment of PA followed two operationalizations, everyday PA, and WHO-PA guideline fulfillment. First, an index of “everyday physical activity” was used to assess everyday habitual PA as an important continuous measure of PA (Andersen et al., 2011). This index was comprised of items of the MoMo Physical Activity Questionnaire assessing time spent in leisurely activity including playing outside, gardening, housework, as well as walking and non-sporting cycling (or other non-motorized vehicles such as inline skates or scooters). Playing outside, gardening, and housework were each assessed by two questions about days per week and minutes per day in which the participants pursued the activity (“On how many days do you usually play outside/garden/work in the household during a week?”, “How long do you on average work/play on one of those days?”). The items walking and cycling were assessed by one item each about the daily distance and time participants walk or cycle (Schmidt et al. 2016). Minutes spent playing outside, walking, cycling, gardening, and housework were then combined in an index that reflected the *daily minutes of habitual everyday PA*. Second, we evaluated the fulfillment of the WHO guideline on PA (World Health Organization, 2010) using a single item (“On how many of the last seven days were you physically active for at least 60 minutes a day?”) included in the MoMo Physical Activity Questionnaire (Schmidt et al. 2016) as PA guideline fulfillment is an important marker for health (Bull et al., 2020). The MoMo Physical Activity Questionnaire containing the beforementioned items has shown sufficient psychometric properties in previous studies (Jekauc et al., 2013).

#### Person-specific Covariates

All person-specific covariates were assessed via questionnaire at wave 3 (pre-pandemic). Only COVID-19 infection was assessed at the post-pandemic timepoint using the question “Have you had a COVID infection so far” (“Yes”, “No”).

##### Sociodemographic variables

Sociodemographic data was collected including age, sex, and socioeconomic variables as well weight and height measured by trained test instructors at the test centers. BMI was calculated and then classified into severe underweight, underweight, normal weight, overweight, and obesity based on percentiles for the German population (Kromeyer-Hauschild et al., 2001). Socioeconomic status (SES) was calculated as an index combing information on education level and occupational status of the parents as well as the equivalized disposable household income.

The education level and occupational status were assessed for both parents and each scored on a scale from 1 to 7 points. The higher scores between both parents were used or, in the case of a single parent household, education and occupational scores from the parent living with the child was used. Equivalized disposable household income was transformed into a score ranging from 1 to 7 points and then summed up with the education and occupation points to form the SES score (range: 3-21). This score was then divided into quintiles based on the whole MoMo wave 3 sample. The lowest quintile (3–12 points) represents a low SES, while the highest quintile (18.6–21 points) represents a high SES. Participants within the middle three quintiles (12.2–20.9 points) were assigned a medium SES (Lampert et al., 2018).

##### Mental Health: Strengths and Difficulties Questionnaire

Behavioral and emotional difficulties were assessed using the German version of the strengths and difficulties (SDQ) questionnaire (parent-report for children aged 4-10 years and self-rated version for adolescents aged 11-17 years). Both versions contain 25 items, five each regarding emotional symptoms, conduct problems, hyperactivity and inattention, peer problems, and prosocial behavior. Each statement is rated on a 3-point Likert scale (“Not true”=0, “Somewhat true”=1, “Certainly true”=2). A total problem score was calculated based on the sum across all problem scales (emotional symptoms, conduct problems, hyperactivity and inattention, and peer relationship problems) leading to a score between 0 and 40 points. Based on national norms (Woerner et al., 2002), the total problem score was categorized into the risk categories inconspicuous (0-12 points), at risk (13-15 points), and conspicuous (16-40 points).

### 1.3 Data Analysis and Statistics

The analysis was conducted using the following software packages in R (R Core Team, 2021). Data wrangling was performed using the ‘*tidyverse*’ packages (Wickham et al., 2019). Random-intercept (RI) Models were calculated with ‘*lme4*’(Bates et al., 2015) and p-values were estimated using ‘*lmerTest*’ (Kuznetsova et al., 2017). Assumptions of the RI models were checked using the ‘*performance*’ R package (Lüdecke et al., 2021). *k*-fold cross validation was performed using ‘*cv*’ package (Fox & Monette, 2023). The R package ‘*ggplot2*’ was used for all visualizations (Wickham, 2016). Participants that provided data at the time points pre-pandemic and post-pandemic were included in the analysis resulting in a longitudinal dataset of 945 individuals. Descriptive analyses and visualizations were conducted to characterize the sample, the outcomes and their distributions across the different measurement time points.

To test the change in HrQoL, ST, everyday PA, and WHO PA days and the potential moderating effect of age on the change of HrQoL, ST, everyday PA, and WHO PA days across the pandemic, mixed-effects models were calculated with measurements (level 1) nested in participants (level 2) to acknowledge the nested structure of the data. RI models were fitted to the data with a RI for the individual, allowing baseline levels of the outcome to vary across individuals and accounting for within-person correlation.

Person-specific time-invariant covariates were entered as fixed predictors. These included sex (female/male), SES (low, moderate, high), any COVID infection until post-pandemic (yes/no), total problem score based on the SDQ at wave 3 and body mass index classification. All covariates were chosen based on previous research linking them independently to the outcomes (Aston, 2018; Nobari et al., 2021).

To test H1, we fitted RI models including the predictors time point (pre-pandemic/post-pandemic), age at wave 3 (centered at the sample mean), and covariates. Age at wave 3 (pre-pandemic) was included to the covariate-adjusted models up to the third polynomial to test for nonlinear effects of age. Polynomial terms were retained based on likelihood ratio test and model fit indices (AIC, BIC). The hypothesis H1 was evaluated based on the simple effect of the predictor time point while adjusting for the effects of covariates.

To test H2, we fitted identical RI models adding the interaction between time point (pre-pandemic/post-pandemic) and age (including its polynomials) to each model. H2 was evaluated based on the statistical significance of this interaction effect. Interpretation of significant interaction effects was eased by data visualization for each model.

The added value of each variable of interest (age at wave 3, time point) and their interaction was assessed by comparing model fit indices (AIC, BIC) of the covariate-adjusted models without these variables of interest to covariate-adjusted models with these variables of interest, as well as to covariate-adjusted models with the interaction terms. The level of statistical significance was set *a priori* to α<0.05.

*k*-fold cross validation was performed to account for capitalization on chance, reduce the risk of overfitting, and improve predictive accuracy of our findings (James et al., 2021). We evaluated the model performance and calculate the average prediction error rate. Initially, data were randomly split into five folds (*k* = 5-fold). One fold was left out (test set), while the four folds were used together to compute the model (training set). Then, the model was tested using the test set. Mean squared error (MSE) was computed on hold-out folds and averaged across folds to quantify the prediction error. Bias-adjusted CV MSEs are reported to approximate the expected prediction error of models fitted on the full sample. This process was repeated until each of the five folds served as the test set. We computed the average of the five MSE values, referred to as the cross-validation error serving as the performance metric for the model of interest. We then compare the MSE between models investigating the same outcome. In an exploratory analysis, we explored the trajectories throughout the lockdowns using repeated contrasts for measurements. This compared pre-pandemic to lockdown1, lockdown1 to lockdown2 and lockdown2 to post-pandemic for the outcome variables controlling for the same covariates as in the models testing H1. Interaction with age was explored similarly to the H2 model.

## Results

### Sample Characteristics

2851 children and adolescents who participated in wave 3 of the MoMo Study prior to the Lockdown I were contacted for participation in the longitudinal assessment. Of those participating, 945 children and adolescents (33.1% response rate, 49% female) had data across both Lockdown I & II and the post-pandemic online assessments and were included in the analysis. Participants were 4-17 years old at the first measurement point (*M*=10.2, *SD*=3.96), and 575 had a medium SES (60.8%), 230 a high SES (24.3%), and 122 a low SES (12.9%). Prevalence of adhd, depression, diabetes, fat metabolism, heart issues, and high blood pressure were 0-2.1% at the first measurement point. See supplement 1 table 1 and table 2 for additional descriptive data of the sample.

**Table 1.** Main results of the linear mixed model for health related quality of life (HrQoL), recreational screen time (ST), everyday physical activity (Everyday PA), and days with >60 minutes moderate-to-vigorous physical activity (WHO PA days) adjusted for age, sex and socioeconomic status, bmi, COVID-19 infection, and risk for mental disorder.

|  | Health-related quality of life |  |  | Recreational screen time |  |  | Everyday physical activity |  |  | Days with ≥60 min of moderate to vigorous physical activity |  |  |
|---|---|---|---|---|---|---|---|---|---|---|---|---|
| *Predictors* | *B* | *CI* | *p* | *B* | *CI* | *p* | *B* | *CI* | *p* | *B* | *CI* | *p* |
| (Intercept) | 55.51 | 54.00 – 57.03 | **<.001** | 179.15 | 162.81 – 195.49 | **<.001** | 116.41 | 105.92 – 126.89 | **<.001** | 4.36 | 4.09 – 4.63 | **<.001** |
| Post-pandemic | -5.92 | -6.82 – -5.03 | **<.001** | 75.17 | 66.65 – 83.69 | **<.001** | -31.15 | -36.40 – -25.91 | **<.001** | -0.27 | -0.43 – -0.11 | **.001** |
| **Number of participants and observations** |  |  |  |  |  |  |  |  |  |  |  |  |
| N | 806<sub>id</sub> |  |  | 818<sub>id</sub> |  |  | 817<sub>id</sub> |  |  | 818<sub>id</sub> |  |  |
| Observations | 1371 |  |  | 1626 |  |  | 1585 |  |  | 1633 |  |  |

**Table 2.**
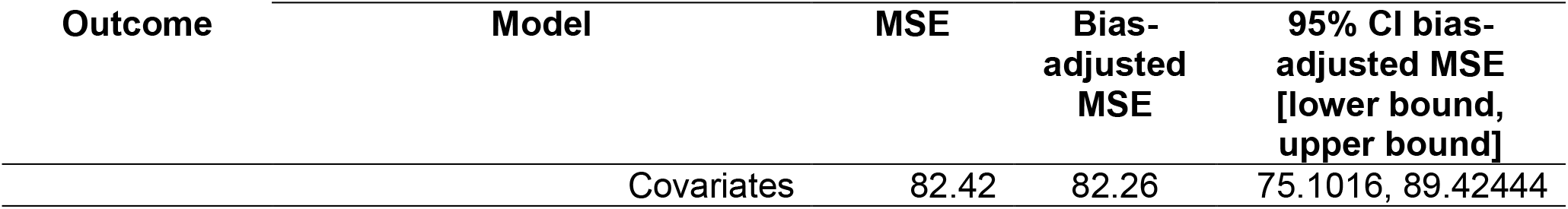
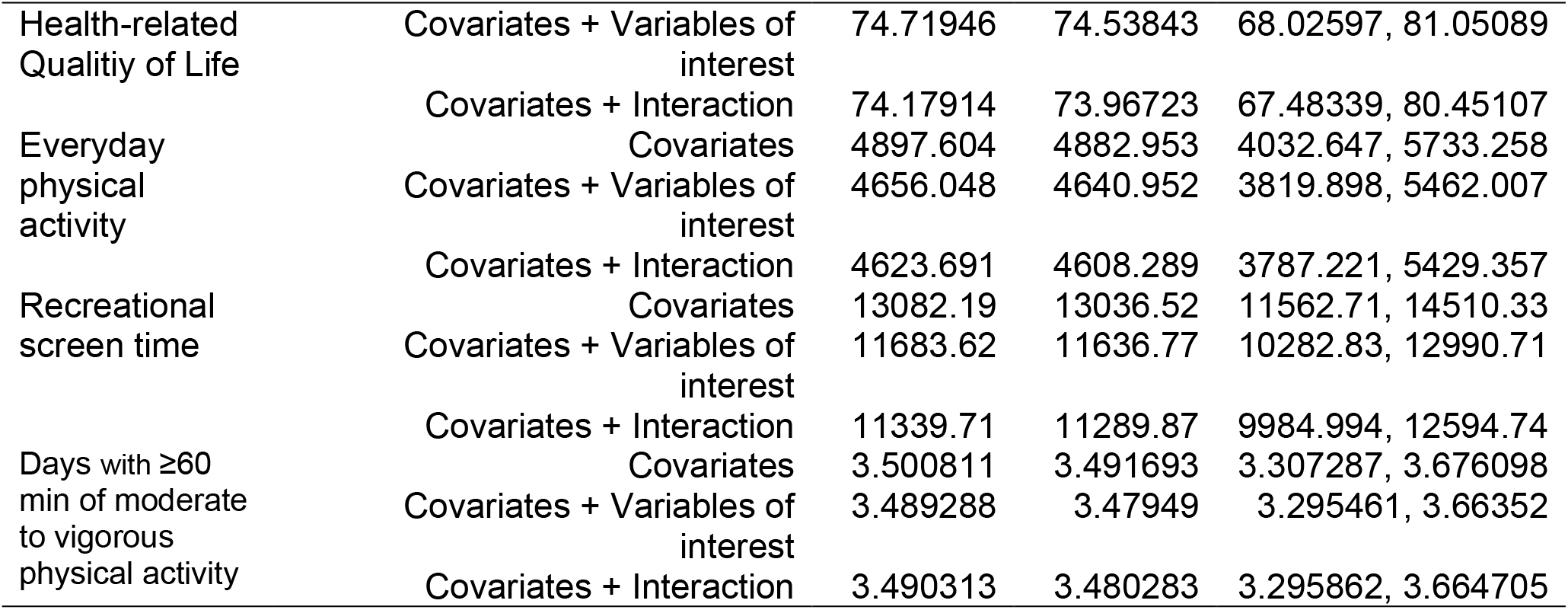
Cross-Validation: Mean Squared Error (MSE) bias-adjusted MSE and 95% confidence intervals of the bias-adjusted MSE for all outcomes across all model groups.

### H1: Change from Pre– to Post-Pandemic

Compared with pre-pandemic levels, post-pandemic levels were significantly lower in lower in HrQoL (b = –5.92, CI [-6.82 – –5.03], p < .001), minutes of everyday PA (b = –31.15, CI [-36.40 –-25.91], p < .001), and WHO PA days (b = –-0.27, CI [-0.43 – –0.11], p = .001), and significantly in higher minutes of ST (b = 75.17, CI [66.65 – 83.69], p < .001) after controlling for covariates (see Table 1, and Supplement 1 Table 4). Cross-validation, R², and model comparison with AIC confirms a better model fit by including our variables of interest compared to the covariate model (see Supplement 1 Table 3).

### H2: Change from Pre– to Post-Pandemic: Moderation by Age

The difference from pre– to post-pandemic is moderated by age and its and its higher-order polynomial terms for all outcome variables except WHO PA days. Full model output is displayed in Supplement 1 Table 5. Data visualization based on age categories indicates that the moderation effect for HrQoL is small, with participants older than 6 years showing a slightly greater decline from pre to post compared to those younger than 6 years (see Supplement 2 Fig. 1). For everyday PA, the moderation effect reveals a greater decline from Pre to Post in children younger than 11 years compared to older children and adolescents (see Supplement 2 Fig. 2). No significant moderation effect was found for WHO PA days (see Supplement 2 Fig. 3). Regarding ST, children and adolescents aged 6–13 years exhibit a greater increase from Pre to Post compared to younger children aged 4–5 years and, in particular, adolescents older than 14 years (see Fig. 1).

**Figure 1.**
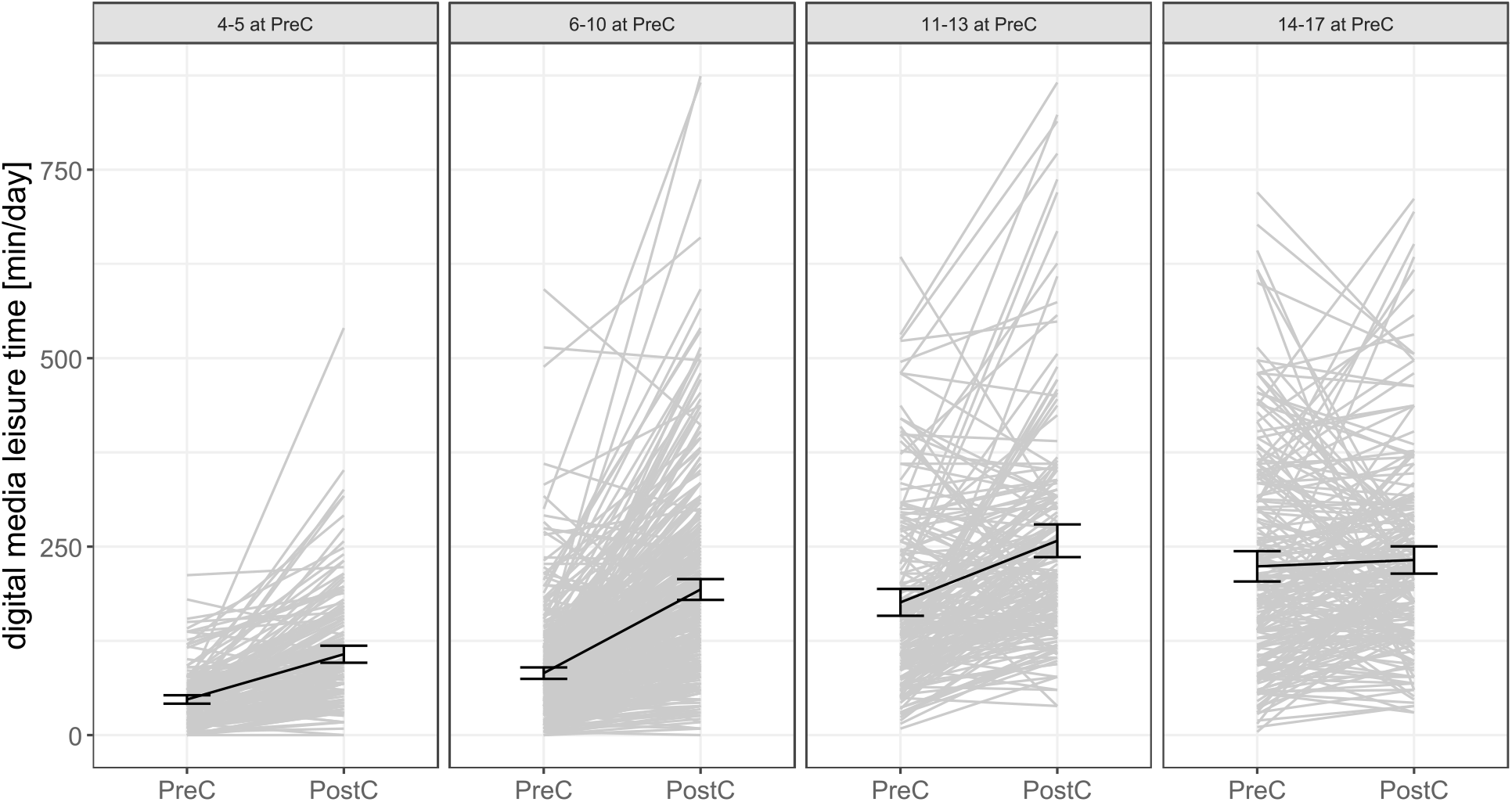
Moderation effect of age pre-pandemic on the differences of digital media use during leisure time from pre-pandemic (PreC) to post-pandemic (PostC). The numbers (e.g., 4-5) indicate the age group at the first measurement PreC. Displayed are individual trajectories, the means, and the 95% Confidence Intervals.

**Figure 2.**
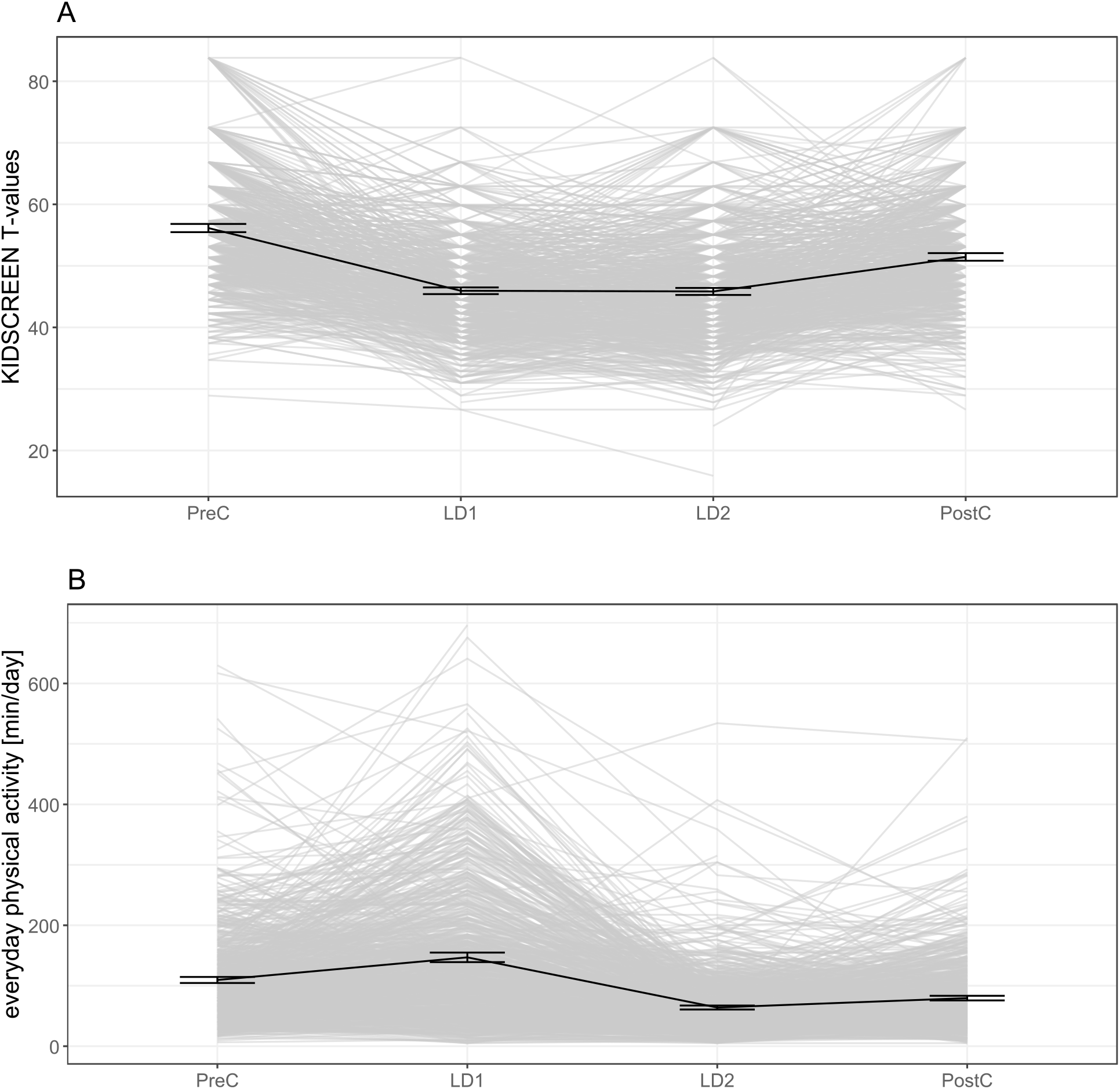
Exploring differences for the variables health related quality of life. (A), and everyday physical activity (B) throughout the four measurements pre-pandemic (PreC) lockdown 1 (LD1), lockdown 2 (LD2), and post-pandemic (PostC). Displayed are individual trajectories, the means, and the 95% Confidence Intervals.

### 3. Cross-Validation

In the cross-validation analysis (see Table 2), adding the variable of interest (VoI) to the covariate-only random-intercept models was associated with lower bias-adjusted mean squared error (MSE) for HrQoL (74.54, 95% CI [68.03-81.05], vs. 82.26, 95% CI [75.10-89.42]), physical activity (PA; 4,640.95, 95% CI [3,819.90-5,462.01], vs. 4,882.95, 95% CI [4,032.65-5,733.26]), and ST (11,636.77, 95% CI [10,282.83-12,990.71], vs. 13,036.52, 95% CI [11,562.71-14,510.33]). Including the interaction term resulted in additional reductions in bias-adjusted MSE for HrQoL (73.97, 95% CI 67.48-80.45), PA (4,608.29, 95% CI 3,787.22-5,429.36), and ST (11,289.87, 95% CI 9,984.99-12,594.74). In contrast, prediction errors for WHO PA days were similar across the covariate-only (3.492, 95% CI 3.307-3.676), covariate-plus-VoI (3.479, 95% CI 3.295-3.664), and interaction (3.480, 95% CI 3.296-3.665) models. Overall, these results suggest modest improvements in predictive performance after inclusion of the VoI and the interaction term for HrQoL, PA, and ST, but only little improvements for WHO PA days. Because the reported confidence intervals pertain to each model’s MSE rather than to paired differences between models, these comparisons should be interpreted descriptively rather than as tests of statistical significance of the different models.

### 4. Exploratory results from Analysis with Contrasts

Exploring the successive differences throughout the two lockdown periods in Germany (see Supplement 1 Table 6) showed lower HrQoL, higher ST, everyday PA and WHO PA days from pre-pandemic to lockdown1 (all *p* < .001). Differences between lockdown1 and lockdown2 indicated higher values of ST, and lower values for everyday PA and WHO PA days (all *p* <.001) with no difference for HrQoL (*p* = .763). Finally, the difference from lockdown2 to post-pandemic showed higher HrQoL, everyday PA, and WHO PA days and lower ST (all *p* < .001). Figure 2 and Supplement 2 Figure 4 visualize the effects and the full model output is displayed in Supplement 1 Table 6.

The described changes in HrQoL, ST, PA, and WHO PA days across time points are moderated by age at wave 3 and its higher-order polynomials from pre-pandemic to lockdown1 for HrQoL, everyday PA, and WHO PA days, from lockdown1 to lockdown2 for ST, and everyday PA, and from lockdown2 to post-pandemic for HrQoL, ST, and everyday PA. The full model output is displayed in Supplement 1 Table 7 and the effects are visualized in Supplement 2 Figures 5, 6, 7, and 8.

## Discussion

This study evaluated the longitudinal change in HrQoL, ST, everyday PA, and WHO PA days in children and adolescents in Germany from pre– to post-pandemic. HrQoL, everyday PA, and the number of days on which the WHO-recommendation for PA was met decreased from pre– to post-pandemic, while ST increased when accounting for other relevant covariates. Age at the beginning of the pandemic moderated the reported change in HrQoL, ST, and everyday PA-but not WHO PA days. Additionally, the changes in these variables throughout the two lockdown periods were explored and showed different patterns of change across HrQoL, ST, PA, and WHO PA days. HrQoL followed a U-shaped change indicating worse HrQoL during the lockdowns and a recovery towards the initial wave 3 levels after restrictions were lifted. ST increased successively throughout the lockdowns with a small reduction at post-pandemic follow-up. Everyday PA and WHO PA days increased in lockdown1, dropped below baseline in lockdown2, and recovered slightly at post-pandemic follow-up. These effects were moderated by age prior to the pandemic with limited moderation effects for WHO PA days.

## Discussion of Study Findings

### H1: Change in HrQoL, ST, PA, and WHO PA days from the pre– to the post-pandemic period

In our sample, HrQoL, PA and ST did not return to pre-pandemic levels within the observed follow-up period. Previous studies found comparable results regarding an increase of ST throughout the pandemic that stayed high afterwards (Liu et al., 2024; Madigan et al., 2022), a decline in HrQoL during the lockdowns that did not recover to the pre-pandemic status (Kopp et al., 2024; Orban et al., 2024; Wolf & Schmitz, 2024), and reduced PA levels in the post-pandemic period (Liu et al., 2024).

The changes in HrQoL, ST, and PA across the COVID-19 pandemic are consistent with previous research. Nevertheless, they may partly reflect normative developmental trends, particularly declining PA and increasing sedentary and screen-based behavior with age (Dumith et al., 2011; Farooq et al., 2020; Pearson et al., 2017). The pandemic may have amplified these developmental patterns by substantially disrupting young people’s daily routines. Systematic reviews indicate that pandemic-related restrictions were associated with increased ST and decreased PA among children and adolescents (Neville et al., 2022; Rossi et al., 2021; Trott et al., 2022). Prolonged reliance on digital media for education, leisure, and social interaction may also have contributed to the normalization and persistence of elevated ST following the easing of restrictions, although longitudinal evidence regarding the durability of these changes remains heterogeneous (Madigan et al., 2022; Neville et al., 2022). Similarly, the closure of schools and sports facilities and the suspension of organized activities reduced opportunities for habitual PA and may have facilitated the development of more sedentary routines (Neville et al., 2022; Rossi et al., 2021). Such patterns may have been reinforced within peer groups, given evidence that adolescents’ PA and sedentary behaviors are associated with those of their friends (Sawka et al., 2013). Persistent reductions in HRQoL may reflect continued pandemic-related uncertainty, family stress, broader psychosocial burdens, increased academic demands, and difficulties readjusting to in-person schooling. This interpretation is consistent with findings from the German COPSY study, which documented substantial reductions in HRQoL and increases in mental health problems during the pandemic, with family climate and socioeconomic disadvantage identified as important correlates (Ravens-Sieberer et al., 2022). Overall, these findings suggest that pandemic-related disruptions may have had sustained consequences for young people’s health behaviors and well-being while simultaneously interacting with normative age-related developmental processes.

### H2: The role of age prior to the pandemic – age-dependent moderation of the change in HrQoL, ST, PA, and WHO PA days from pre-to post-pandemic period

In our sample, age moderated the change from pre-to post-pandemic indicating that changes in HrQoL, ST, and PA differed across developmental stages prior to the onset of the pandemic.

Children aged 6-13 years appeared particularly vulnerable to reductions in HRQoL. This developmental period encompasses school-related transitions and increasing reliance on peer relationships, which are central to social and emotional development. Consequently, restricted peer contact and loneliness may have been especially detrimental to well-being (Loades et al., 2020; Orben et al., 2020; Ravens-Sieberer et al., 2022). Visual inspection of age group-specific changes in ST suggested that ST increased particularly from approximately six years of age, coinciding with school entry and expanding access to digital devices, whereas ST remained consistently high in the oldest age group. This pattern is consistent with evidence that screen-based and sedentary behaviors increase across childhood and adolescence, although the precise age-related trajectories vary by screen activity and context (Pearson et al., 2017; Thomas et al., 2020). Age-specific peculiarities differed from previous studies as the meta-analysis of (Madigan et al., 2022) found the largest effect for individuals aged 12 to 18 years while our results show the largest effects from 6-13-year-olds. Younger children showed more pronounced deteriorations in PA, potentially because their activity is more dependent on structured opportunities, school environments, and parental support than that of older children (Neville et al., 2022; Rossi et al., 2021). By contrast, the number of days on which participants met the WHO PA recommendation appeared comparatively stable. This may partly reflect the limited sensitivity of a categorical measure: because the WHO recommendation defines sufficient activity as at least 60 minutes of moderate-to-vigorous PA per day, substantial variation within the range of 0–59 minutes may not produce a change in classification (World Health Organization, 2010). Our findings suggest that age is an important moderator of health-related life aspects in youth in the face of adversity.

Cross-validation adds an important distinction between detecting associations and improving prediction. Including baseline age and measurement time point reduced prediction error for HRQoL, everyday PA, and ST, suggesting that these variables captured information beyond the included covariates. However, adding the age-by-time interactions produced only small further improvements. Thus, although age-related differences in change were statistically detectable, their additional contribution to prediction was limited. These findings support considering developmental stage when interpreting population-level patterns, but caution against treating age as a sufficient basis for individual risk identification. For WHO PA days, the negligible improvement in prediction indicates that the added variables contributed little predictive information, despite the statistically significant average pre–post decline. This may partly reflect the threshold-based measure, which does not capture changes in activity duration within days above or below the 60-minute threshold. Overall, cross-validation provides a complementary assessment of predictive performance, rather than independent confirmation of the proposed mechanisms or the causal role of the pandemic. The differences in prediction error should also be interpreted descriptively, because uncertainty was reported for each model’s error rather than for paired differences between models.

### Exploratory analysis of the trajectory throughout the lockdown periods

The exploratory analysis shows that different aspects of health follow different recovery patterns throughout the four measurement time points underscoring the benefit of higher resolution longitudinal assessments. The exploratory trajectories suggest that recovery following the pandemic was uneven across health domains rather than a uniform return to pre-pandemic conditions. HRQoL improved after the lockdown periods but remained below baseline, whereas ST remained elevated. Everyday PA and WHO PA days showed a different pattern, with an initial increase followed by a decline and only partial recovery. This divergence suggests that psychological well-being and health behaviors may respond differently to the introduction and removal of restrictions. Improvements in well-being following reopening did not necessarily coincide with a return to earlier activity and media-use patterns in this observational study. The initial increase in PA is particularly relevant because it challenges an interpretation of restrictions as uniformly detrimental to activity. One possible explanation is that some young people initially compensated for disrupted routines through informal activity, but that these opportunities or adaptations were not maintained during the subsequent pandemic phase. Contextual differences between the two lockdowns may have contributed to this pattern: the first lockdown coincided with considerably better weather than the second, which likely favored outdoor activity, and families may have had more discretionary time, as digital options for home office and remote schooling were less established than during the second lockdown. Similarly, persistently elevated ST is compatible with the establishment of routines that continued after restrictions ended. Similarly, persistently elevated ST is compatible with the establishment of routines that continued after restrictions ended. These explanations remain tentative, as the relevant mechanisms were not known. Taken together, the findings suggest that restoring access to schools, social contact, and organized activities may not by itself be sufficient to restore pre-pandemic health behaviors. Recovery may require domain-specific support, with attention to the developmental differences indicated by the age interactions. Because these analyses were exploratory and lacked an unexposed comparison group, the trajectories should be interpreted as patterns observed across the pandemic period rather than as isolated effects of particular restrictions.

### Implications of the Study Findings

Collectively, these findings suggest that developmental transition periods may represent critical windows during which changes in daily routines and social environments have implications for health behavior and well-being (Blakemore & Mills, 2014). This interpretation may be particularly relevant in the context of ongoing global crises affecting youth’s everyday lives, such as financial crisis and concerns about war. Young people of similar ages may nevertheless follow substantially different trajectories, and identifying those most in need of support is likely to require information beyond the predictors included here. Additionally, HRQoL, ST, and PA indicators may not be independent outcomes. Digital media, for example, may have increased sedentary time and reduced PA opportunities while simultaneously compensating for restricted in-person contact by facilitating social connection. Conversely, excessive or maladaptive use may have adversely affected well-being (Odgers & Jensen, 2020). Although the present study examined these outcomes separately, they may influence one another over time-both within and between individuals. It would be interesting to better investigate their reciprocal longitudinal associations in future research.

### Strengths and Limitations

This study has both strengths and limitations. The main strength is the longitudinal sample of nearly 1,000 participants across a wide age range, combined with a study design featuring multiple measurements at relevant time points: a proximal assessment pre-pandemic, two assessments during lockdowns, and a post-pandemic assessment after relevant restrictions were lifted. Furthermore, health is operationalized as health behavior (ST, everyday PA, and WHO PA days) supplemented by a psychological measure (HrQoL) within a single sample, enabling the comparison of trajectories in these parameters over time. Lastly, PA is assessed using two different constructs: everyday PA captures important facets of children’s and adolescents’ everyday lives, and WHO PA days focus on PA of at least moderate intensity with a clear health-beneficial cut-off.

The main limitation of this study is that no control group was present, and the longitudinal design is therefore confounded by the participants’ increasing age; the effects of COVID and the passage of time cannot be clearly differentiated with this approach. Furthermore, the direct comparison of developmental changes with pre-pandemic findings is limited, as comparable studies with similar designs (i.e., temporal spacing and age range) are lacking. However, as discussed earlier, the patterns of change across the four measurement points clearly differ from cross-sectionally reported differences between age groups (Schmidt et al. under review), which highlights the importance of this contribution. A second limitation is that the present study relies on a non-random subsample of a nationally representative sample and is therefore no longer representative of the general population. Nonetheless, the large longitudinal sample, together with a cross-validation approach that accounts for capitalization on chance, reduces the risk of overfitting, and improves predictive accuracy, strengthens the generalizability of our findings. Third, the change in assessment mode from in-person questionnaires at the pre-pandemic assessment to online questionnaires at subsequent assessments may have introduced method-related measurement effects. As assessment mode was confounded with measurement time point, such effects cannot be disentangled from the observed longitudinal changes. Finally, all outcomes are based on subjective self-report assessments. Although all instruments were validated, future studies should combine self-report with objective measures, employing sensor-based PA assessment (Burchartz et al., 2020; Fiedler et al., under review), technology based tools for ST (Perez et al., 2023), or ecologically momentary assessment (Reichert et al., 2020) approaches for higher-resolution data with greater ecological validity.

### Conclusion

Our findings that trajectories of HrQoL, ST, and PA throughout the COVID-19 pandemic are strongly related to pre-pandemic age, emphasize the need for continuous, developmentally tailored support, particularly during critical transitions such as school entry and the move to secondary education. Routine monitoring of HrQoL, ST, and PA in schools and pediatric care may enable early identification and prevention of adverse trajectories in the case of crises or critical changes in youth’s everyday lives. During online schooling, children, parents, and teachers should receive guidance on balancing screen exposure with movement, recovery, and social interaction (Bull et al., 2020; Odgers & Jensen, 2020). Consistent with the behavioral and cultural insights approach, interventions should extend beyond individual responsibility by ensuring opportunities to develop movement competence, participate in PA, and maintain peer relationships (World Health Organization, 2022).

## Declarations

### Author contribution

Conceptualization: CL, JF; Methodology: CL, JF; Data curation and formal analysis: CL, JF, LK; Writing – original draft preparation: CL, JF; Writing – review and editing: CL, JF, LK, CV, CN, MV, AW; Funding acquisition: AW, CN; Project Administration: CN

## Data Availability

All data produced in the present study are available upon reasonable request to the authors

## Acknowledgements

We would like to express our sincere gratitude to the dedicated student assistants and test instructors who contributed significantly to the successful data collection of this study. Without their dedicated support and tireless work, this research would not have been possible.

