## Supplement_2_Figures for "Longitudinal changes of health-related quality of life, digital media use, and physical activity in children between 4-17 years during the COVID-19 pandemic in Germany"

Supplement Figure 1. Moderation effect of age pre-pandemic on the differences of health related quality of life from pre-pandemic (PreC) to post-pandemic (PostC). The numbers e.g. 4-5 indicate the age group at the first measurement PreC.

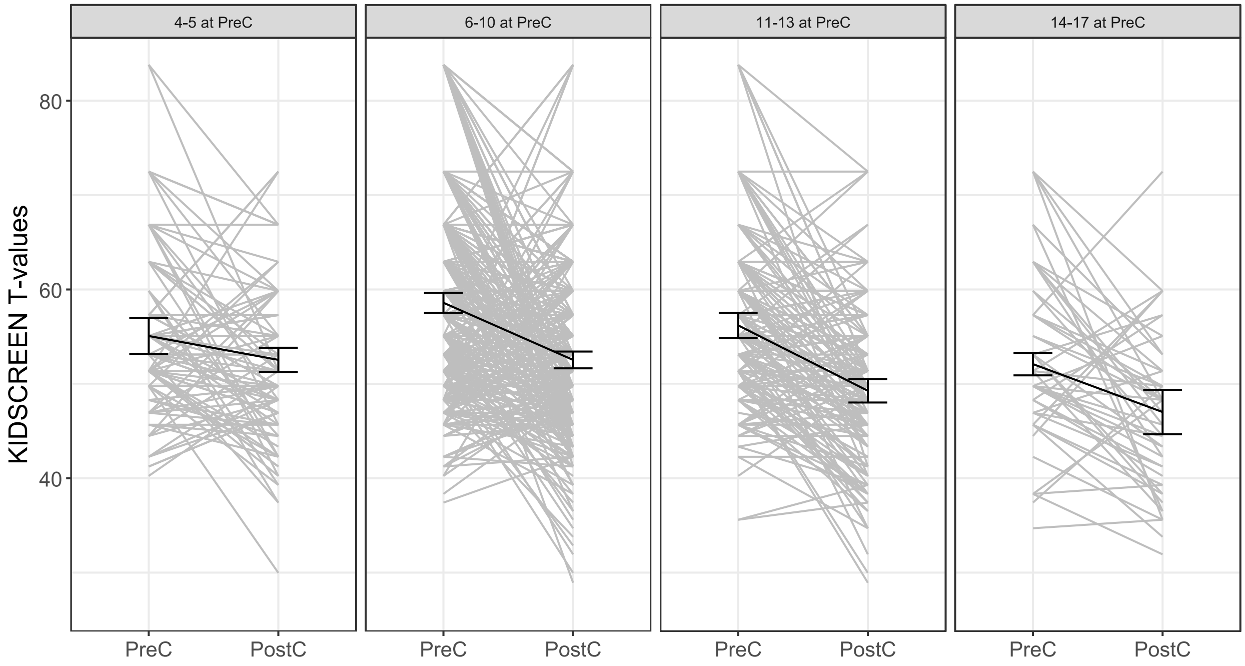

Supplement Figure 2. Moderation effect of age pre-pandemic on the differences of everyday physical activity from pre-pandemic (PreC) to post-pandemic (PostC). The numbers e.g. 4-5 indicate the age group at the first measurement PreC.

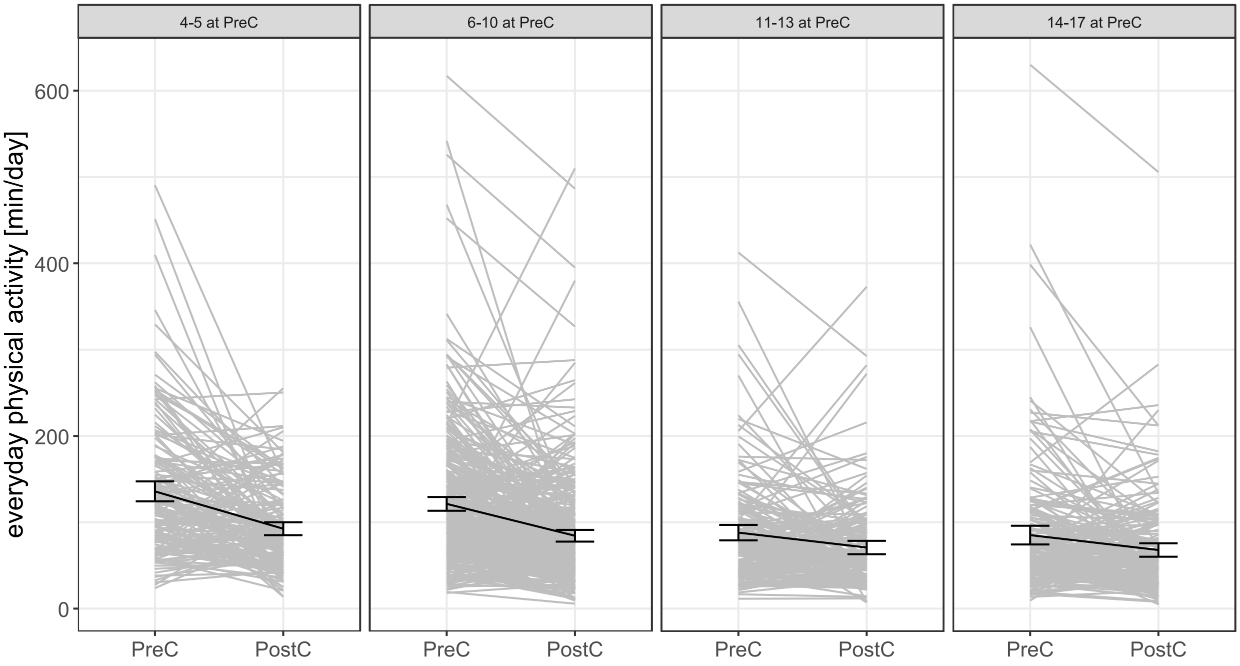

Supplement Figure 3. Moderation effect of age pre-pandemic on the differences of days with more than 60 minutes moderate-to-vigorous physical activity (MVPA) from pre-pandemic (PreC) to post-pandemic (PostC). The numbers e.g. 4-5 indicate the age group at the first measurement PreC.

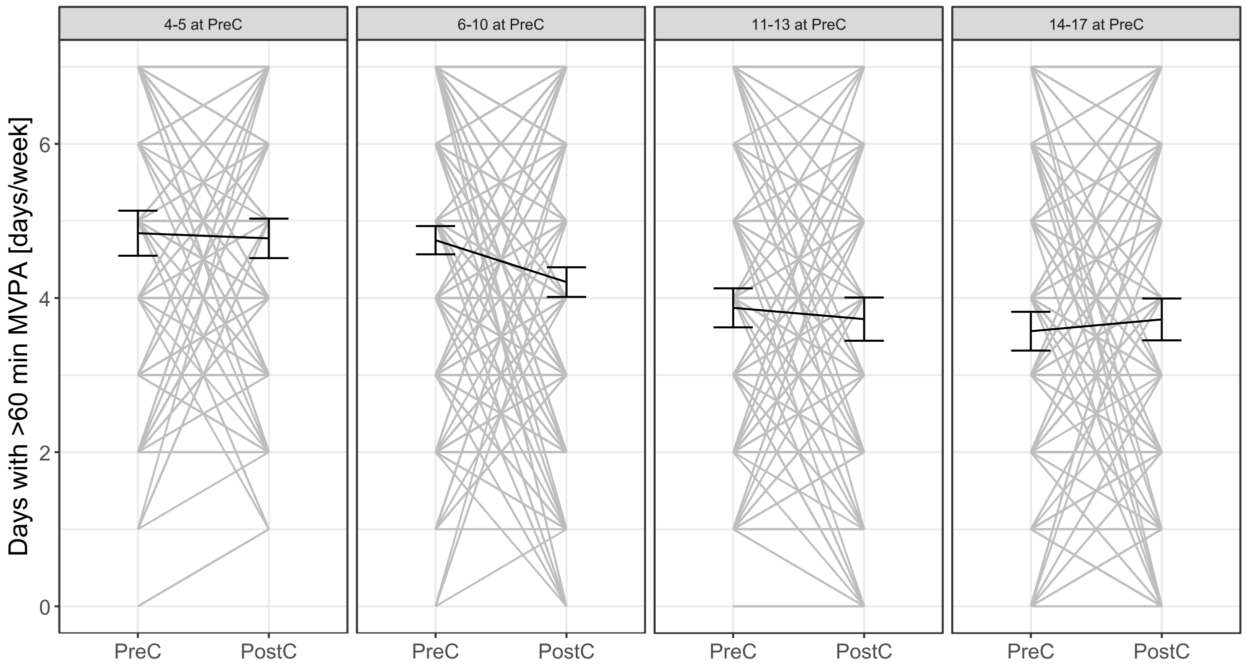

Supplement Figure 4. Exploring differences for the variables digital media use during leisure time (A), and days with more than 60 minutes moderate-to-vigorous physical activity (MVPA) (B) throughout the four measurements pre-pandemic (PreC) lockdown 1 (LD1), lockdown 2 (LD2), and post-pandemic (PostC).

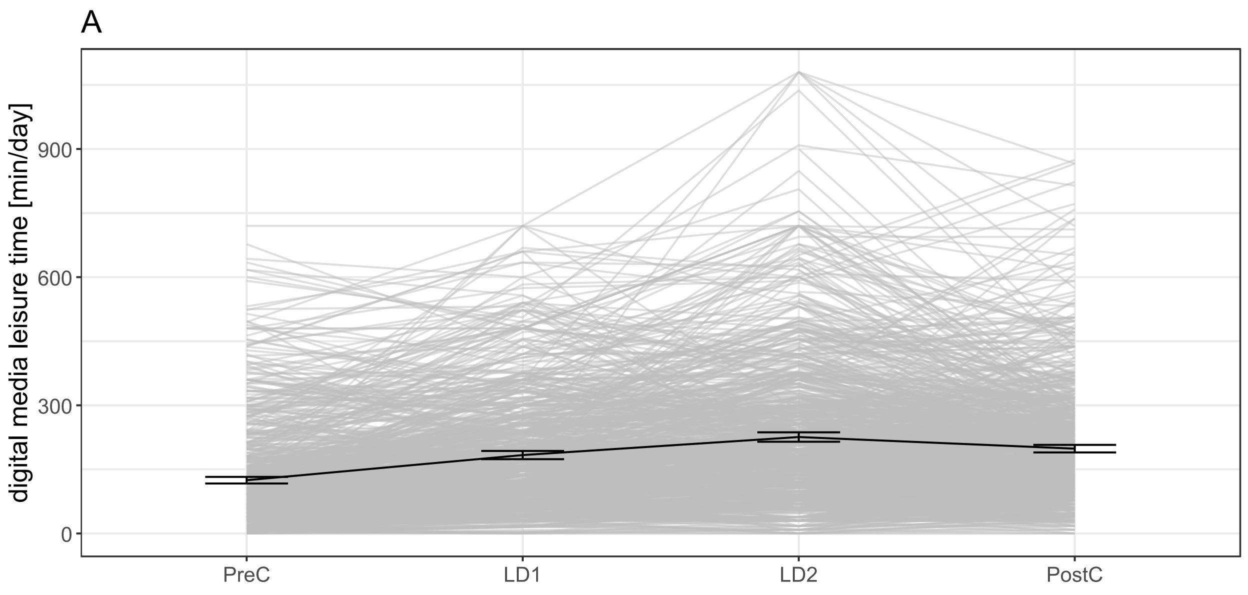


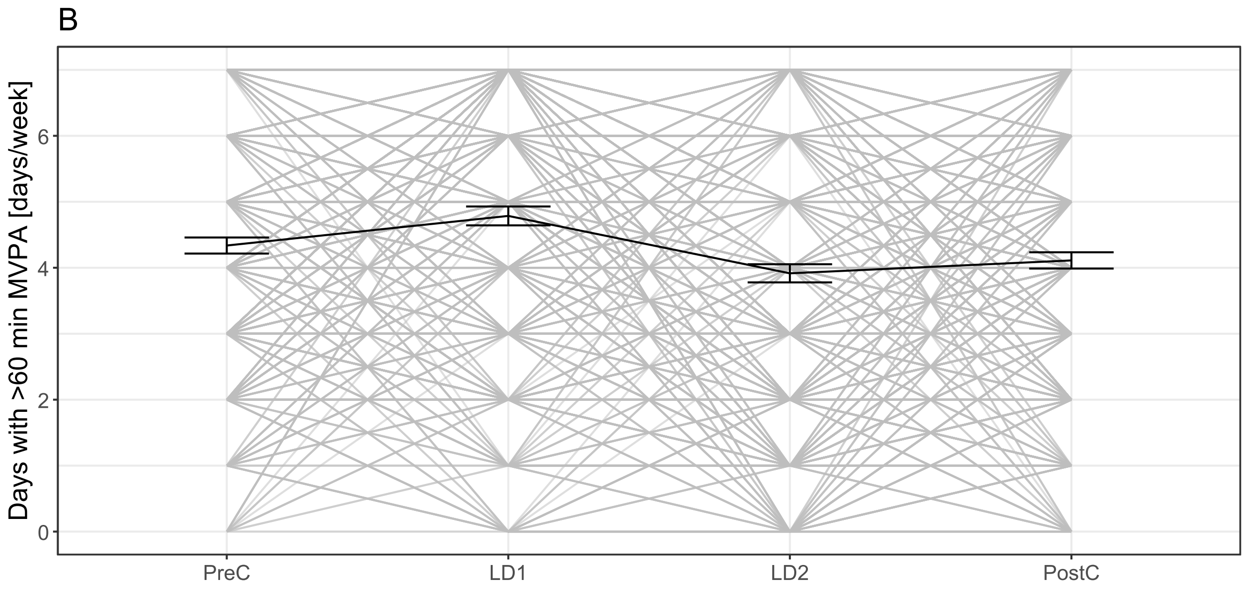

Supplement Figure 5. Moderation effect of age pre covid on the differences of health related quality of life throughout the four measurements pre-pandemic (PreC) lockdown 1 (LD1), lockdown 2 (LD2), and post-pandemic (PostC). The numbers e.g. 4-5 indicate the age group at the first measurement PreC.

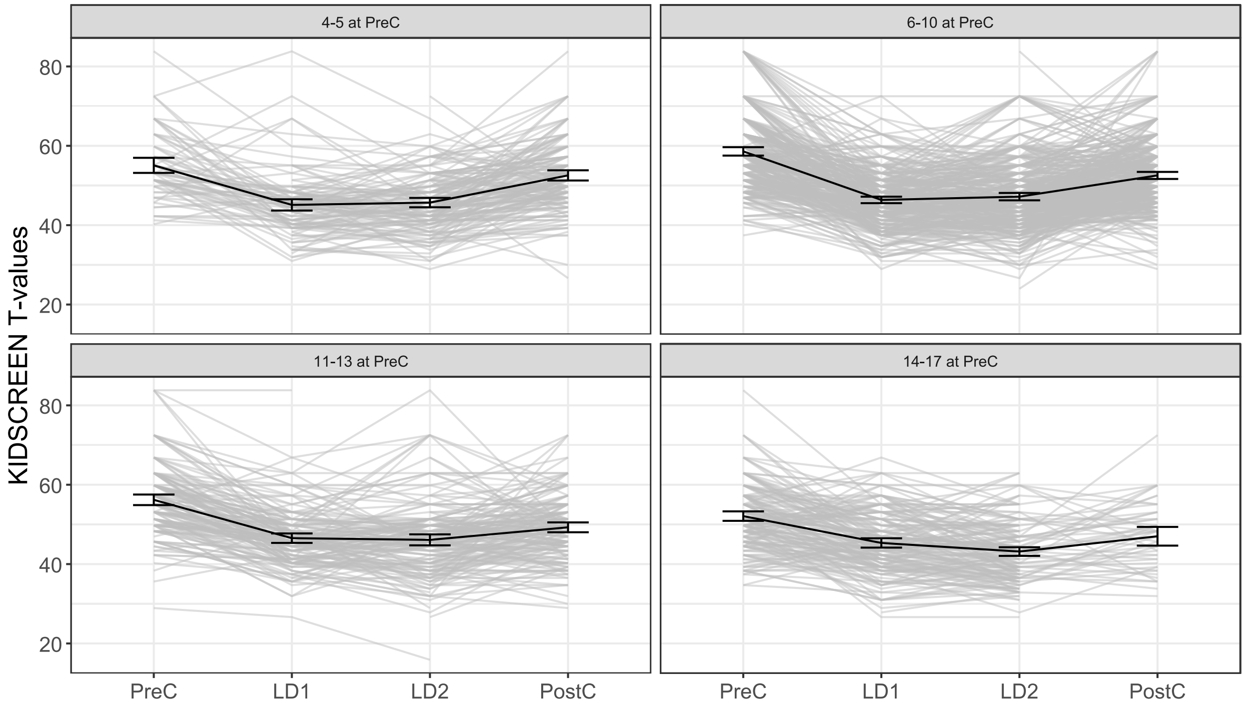

Supplement Figure 6. Moderation effect of age pre covid on the differences of digital media leisure time throughout the four measurements pre-pandemic (PreC) lockdown 1 (LD1), lockdown 2 (LD2), and post-pandemic (PostC). The numbers e.g. 4-5 indicate the age group at the first measurement PreC.

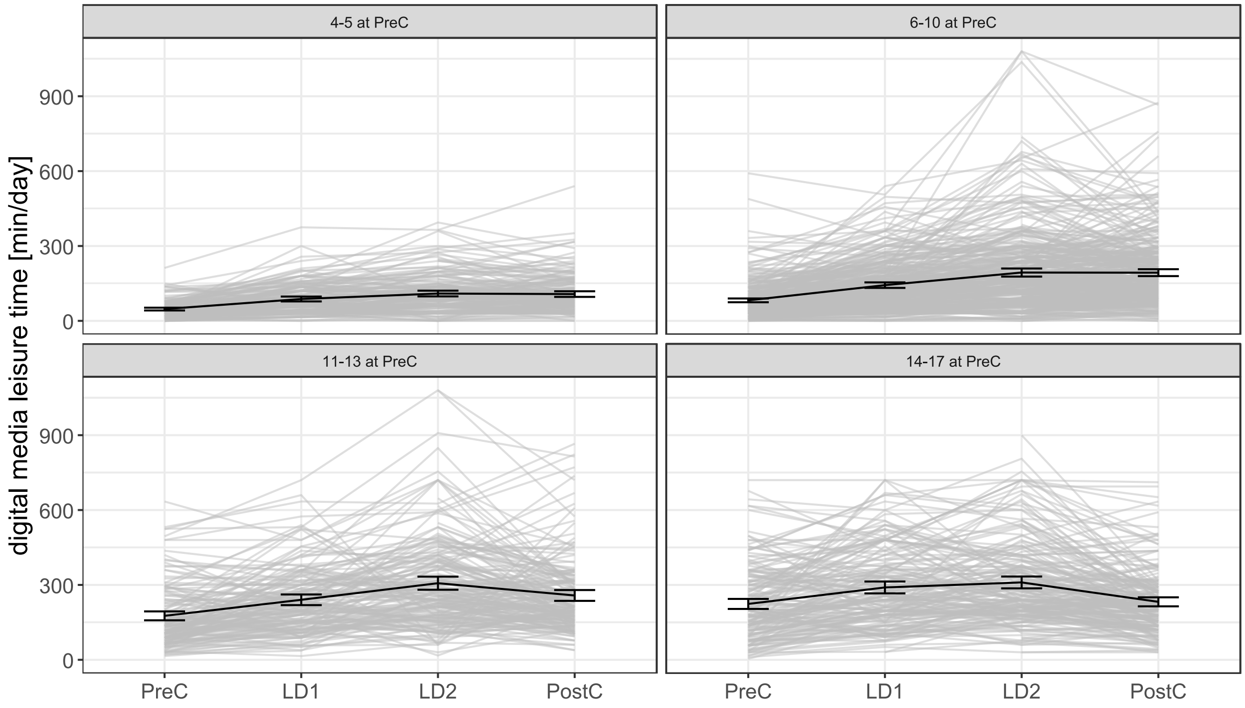

Supplement Figure 7. Moderation effect of age pre covid on the differences of everyday physical activity throughout the four measurements pre-pandemic (PreC) lockdown 1 (LD1), lockdown 2 (LD2), and post-pandemic (PostC). The numbers e.g. 4-5 indicate the age group at the first measurement PreC.

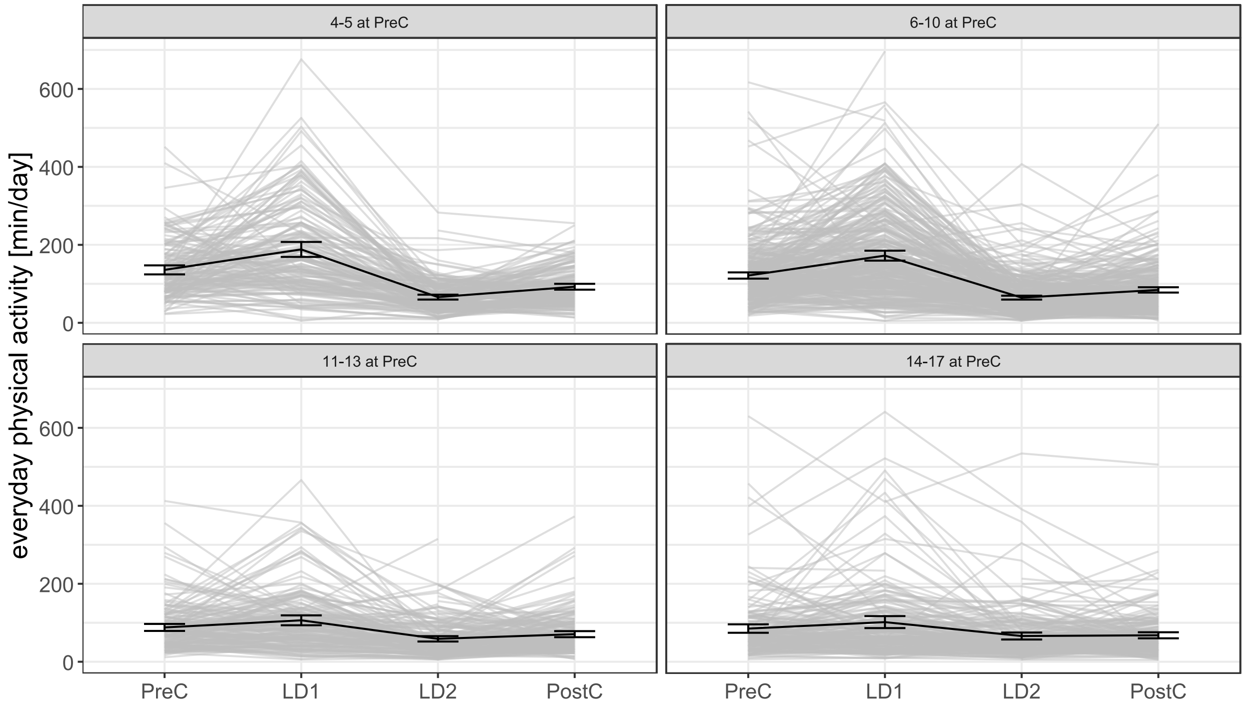

Supplement Figure 8. Moderation effect of age pre covid on the differences of days with more than 60 minutes moderate-to-vigorous physical activity (MVPA) throughout the four measurements pre-pandemic (PreC) lockdown 1 (LD1), lockdown 2 (LD2), and post-pandemic (PostC). The numbers e.g. 4-5 indicate the age group at the first measurement PreC.

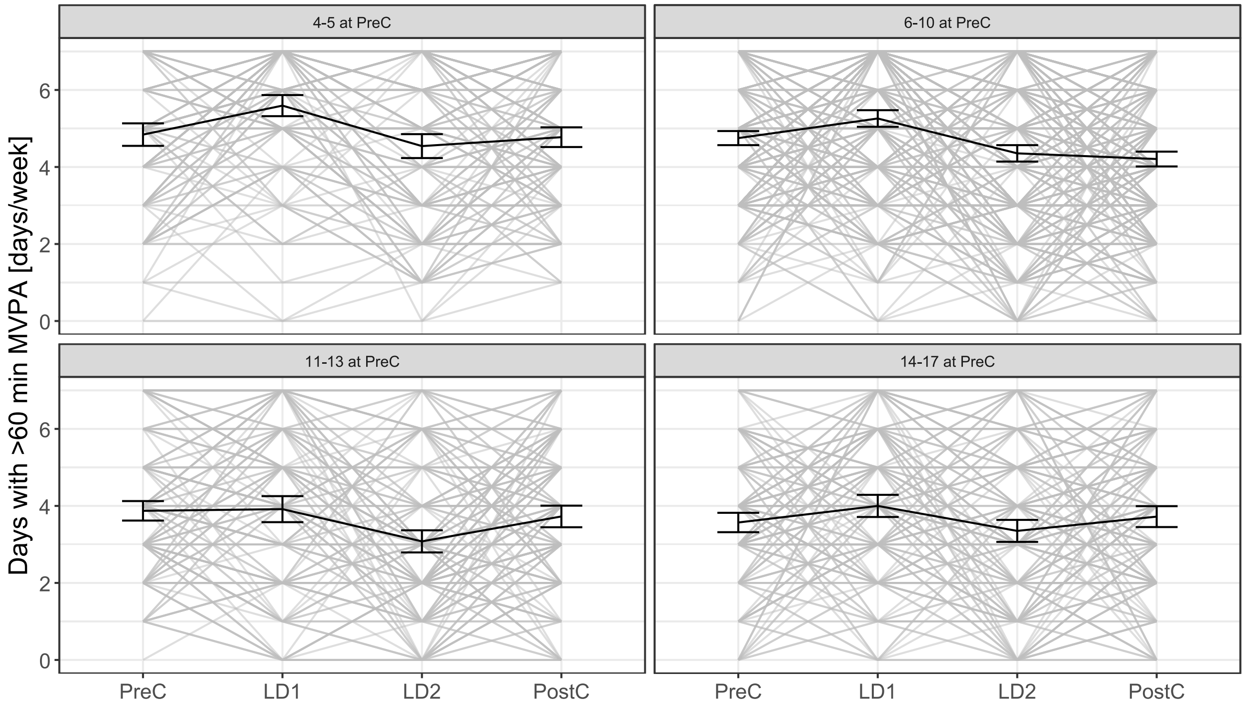
