## Supplement_1_Tables for "Longitudinal changes of health-related quality of life, digital media use, and physical activity in children between 4-17 years during the COVID-19 pandemic in Germany"

### Supplemental Material

Supplement Table 1. Participant characteristics prior to the pandemic (wave 3, 2018-2020).

| | **male (N=467)** | **female (N=478)** |
|---|---|---|
| **age** | | |
| Mean (SD) | 9.79 (3.84) | 10.6 (4.04) |
| **Socioeconomic status** | | |
| medium | 278 (59.5%) | 297 (62.1%) |
| low | 58 (12.4%) | 64 (13.4%) |
| high | 122 (26.1%) | 108 (22.6%) |
| Missing | 9 (1.9%) | 9 (1.9%) |
| **BMI** | | |
| normal | 354 (75.8%) | 342 (71.5%) |
| severe underweight | 9 (1.9%) | 18 (3.8%) |
| underweight | 26 (5.6%) | 24 (5.0%) |
| overweight | 20 (4.3%) | 24 (5.0%) |
| obesity | 10 (2.1%) | 12 (2.5%) |
| Missing | 48 (10.3%) | 58 (12.1%) |
| **covid infection** | | |
| no | 136 (29.1%) | 173 (36.2%) |
| yes | 327 (70.0%) | 303 (63.4%) |
| Missing | 4 (0.9%) | 2 (0.4%) |
| **risk for mental disorder** | | |
| inconspicuous | 365 (78.2%) | 374 (78.2%) |
| at risk | 44 (9.4%) | 64 (13.4%) |
| conspicuous | 53 (11.3%) | 36 (7.5%) |
| Missing | 5 (1.1%) | 4 (0.8%) |
| **adhs** | | |
| no adhs | 449 (96.1%) | 462 (96.7%) |
| adhs | 10 (2.1%) | 4 (0.8%) |
| Missing | 8 (1.7%) | 12 (2.5%) |
| **depression** | | |
| no depression | 458 (98.1%) | 461 (96.4%) |
| depression | 2 (0.4%) | 6 (1.3%) |
| Missing | 7 (1.5%) | 11 (2.3%) |
| **diabetes** | | |
| no | 457 (97.9%) | 464 (97.1%) |

| | male (N=467) | female (N=478) |
|---|---|---|
| yes | 4 (0.9%) | 0 (0%) |
| Missing | 6 (1.3%) | 14 (2.9%) |
| **lipid metabolism disorder** | | |
| no | 457 (97.9%) | 467 (97.7%) |
| yes | 1 (0.2%) | 0 (0%) |
| Missing | 9 (1.9%) | 11 (2.3%) |
| **heart issues** | | |
| no | 452 (96.8%) | 463 (96.9%) |
| yes | 7 (1.5%) | 1 (0.2%) |
| Missing | 8 (1.7%) | 14 (2.9%) |
| **high blood pressure** | | |
| no | 461 (98.7%) | 462 (96.7%) |
| yes | 0 (0%) | 3 (0.6%) |
| Missing | 6 (1.3%) | 13 (2.7%) |

**Supplement Table 2.** Descriptive Statistics of the outcomes.

| | Pre-pandemic | | Post-pandemic | |
|---|---|---|---|---|
| | **male (N=467)** | **female (N=478)** | **male (N=467)** | **female (N=478)** |
| **Health-related quality of life** | | | | |
| Mean (SD) | 57.0 (9.12) | 55.3 (10.1) | 52.2 (8.53) | 50.6 (8.87) |
| Missing | 69 (14.8%) | 71 (14.9%) | 71 (15.2%) | 108 (22.6%) |
| **Recreational screen time** | | | | |
| Mean (SD) | 130 (124) | 119 (116) | 217 (158) | 180 (112) |
| Missing | 7 (1.5%) | 10 (2.1%) | 4 (0.9%) | 2 (0.4%) |
| **Everyday physical activity** | | | | |
| Mean (SD) | 117 (79.1) | 102 (75.2) | 85.1 (63.0) | 74.2 (53.5) |
| Missing | 9 (1.9%) | 7 (1.5%) | 28 (6.0%) | 18 (3.8%) |
| **Days with ≥60 min of moderate to vigorous physical activity** | | | | |
| Mean (SD) | 4.38 (1.95) | 4.30 (1.85) | 4.29 (1.92) | 3.94 (1.92) |
| Missing | 2 (0.4%) | 5 (1.0%) | 0 (0%) | 0 (0%) |

**Supplement Table 3.** Linear mixed model of health related quality of life, recreational screen time, everyday physical activity, and days with ≥60 minutes moderate-to-vigorous physical activity including only the covariates age centered (and polynomials), sex, socioeconomic status (SES), body mass index (BMI), covid infection, and risk for mental disorder (SDQ) (and polynomials).

| | **Health-related quality of life** | | | **Recreational screen time** | | | **Everyday physical activity** | | | **Days with ≥60 min of moderate to vigorous physical activity** | | |
|---|---|---|---|---|---|---|---|---|---|---|---|---|
| *Predictors* | *Estimates* | *CI* | *p* | *Estimates* | *CI* | *p* | *Estimates* | *CI* | *p* | *Estimates* | *CI* | *p* |
| (Intercept) | 51.78 | 50.50 – 53.06 | **<0.001** | 216.99 | 201.23 – 232.76 | **<0.001** | 101.26 | 91.08 – 111.45 | **<0.001** | 4.22 | 3.97 – 4.48 | **<0.001** |
| Age | -3.07 | -4.26 – -1.89 | **<0.001** | 80.43 | 65.72 – 95.14 | **<0.001** | -4.09 | -5.12 – -3.05 | **<0.001** | -0.11 | -0.13 – -0.08 | **<0.001** |
| Age² | -1.26 | -1.86 – -0.66 | **<0.001** | -10.62 | -17.81 – -3.44 | **0.004** | | | | | | |
| Age³ | 2.39 | 1.03 – 3.75 | **0.001** | -26.56 | -42.32 – -10.80 | **0.001** | | | | | | |
| Sex [male]: ref. | | | | | | | | | | | | |
| Sex [female] | -1.36 | -2.34 – -0.39 | **0.006** | -38.24 | -50.35 – -26.13 | **<0.001** | -8.85 | -16.78 – -0.93 | **0.029** | -0.12 | -0.32 – 0.08 | 0.252 |

| | | | | | | | | | | | | |
|---|---|---|---|---|---|---|---|---|---|---|---|---|
| SES [low] | 0.16 | -1.37 – 1.70 | 0.834 | 40.22 | 21.55 – 58.88 | **<0.001** | 11.72 | -0.50 – 23.93 | 0.060 | -0.08 | -0.39 – 0.22 | 0.589 |
| SES [medium]: ref. | | | | | | | | | | | | |
| SES [high] | 1.99 | 0.86 – 3.11 | **0.001** | -36.72 | -50.90 – -22.55 | **<0.001** | -2.84 | -12.05 – 6.37 | 0.545 | 0.33 | 0.10 – 0.57 | **0.005** |
| BMI [severe underweight] | 1.29 | -1.44 – 4.01 | 0.354 | -3.86 | -37.76 – 30.04 | 0.823 | -7.15 | -29.08 – 14.78 | 0.523 | -0.34 | -0.90 – 0.21 | 0.225 |
| BMI [underweight] | 1.68 | -0.30 – 3.66 | 0.096 | -11.98 | -37.81 – 13.86 | 0.363 | -15.10 | -31.78 – 1.59 | 0.076 | 0.12 | -0.30 – 0.55 | 0.570 |
| BMI [normalweight]: ref. | | | | | | | | | | | | |
| BMI [overweight] | -2.44 | -4.68 – -0.21 | **0.032** | 14.10 | -12.96 – 41.16 | 0.307 | 10.29 | -7.63 – 28.21 | 0.260 | -0.06 | -0.51 – 0.38 | 0.780 |
| BMI [obese] | 1.00 | -1.96 – 3.97 | 0.507 | 28.15 | -9.65 – 65.96 | 0.144 | -14.28 | -39.05 – 10.48 | 0.258 | -0.25 | -0.86 – 0.37 | 0.437 |

| | | | | | | | | | | | | |
|---|---|---|---|---|---|---|---|---|---|---|---|---|
| Covid infection [yes] | -0.60 | -1.72 – 0.53 | 0.298 | -10.36 | -24.18 – 3.46 | 0.142 | -0.33 | -9.11 – 8.45 | 0.942 | -0.18 | -0.40 – 0.04 | 0.118 |
| Covid infection [no]: ref. | | | | | | | | | | | | |
| SDQ | -5.04 | -6.19 – -3.89 | **<0.001** | 42.15 | 27.44 – 56.86 | **<0.001** | -5.10 | -14.64 – 4.43 | 0.294 | -0.35 | -0.59 – -0.11 | **0.005** |
| $SDQ^2$ | 0.50 | -0.84 – 1.85 | 0.463 | -16.88 | -33.40 – -0.36 | **0.045** | -8.64 | -19.43 – 2.15 | 0.116 | -0.15 | -0.42 – 0.13 | 0.293 |
| **Random Effects** | | | | | | | | | | | | |
| $\sigma^2$ | 81.66 | | | 10474.84 | | | 3275.75 | | | 2.76 | | |
| $\tau_{00}$ | 0.21 $_{id}$ | | | 2331.97 $_{id}$ | | | 1544.00 $_{id}$ | | | 0.70 $_{id}$ | | |
| ICC | 0.00 | | | 0.18 | | | 0.32 | | | 0.20 | | |
| N | 806 $_{id}$ | | | 818 $_{id}$ | | | 817 $_{id}$ | | | 818 $_{id}$ | | |
| Observations | 1371 | | | 1626 | | | 1585 | | | 1633 | | |
| Marginal $R^2$ / Conditional $R^2$ | 0.112 / 0.114 | | | 0.282 / 0.413 | | | 0.063 / 0.363 | | | 0.067 / 0.255 | | |

Supplement Table 4. Linear mixed model H1 for health related quality of life, recreational screen time, everyday physical activity, and days with >60 minutes moderate-to-vigorous physical activity adjusted for age centered (and polynomials), sex, socioeconomic status (SES), body mass index (BMI), covid infection, and risk for mental disorder (SDQ) (and polynomials).

| | **Health-related quality of life** | | | **Recreational screen time** | | | **Everyday physical activity** | | | **Days with ≥60 min of moderate to vigorous physical activity** | | |
|---|---|---|---|---|---|---|---|---|---|---|---|---|
| *Predictors* | *Estimates* | *CI* | *p* | *Estimates* | *CI* | *p* | *Estimates* | *CI* | *p* | *Estimates* | *CI* | *p* |
| (Intercept) | 55.51 | 54.00 – 57.03 | **<0.001** | 179.15 | 162.81 – 195.49 | **<0.001** | 116.41 | 105.92 – 126.89 | **<0.001** | 4.36 | 4.09 – 4.63 | **<0.001** |
| Wave [pre-pandemic]: ref. | | | | | | | | | | | | |
| wave [post pandemic] | -5.92 | -6.82 – -5.03 | **<0.001** | 75.17 | 66.65 – 83.69 | **<0.001** | -31.15 | -36.40 – -25.91 | **<0.001** | -0.27 | -0.43 – -0.11 | **0.001** |
| Age | -0.80 | -1.10 – -0.50 | **<0.001** | 79.89 | 65.19 – 94.60 | **<0.001** | -4.05 | -5.09 – -3.02 | **<0.001** | -0.11 | -0.13 – -0.08 | **<0.001** |
| $Age^2$ | -0.09 | -0.13 – -0.05 | **<0.001** | -10.77 | -17.95 – -3.59 | **0.003** | | | | | | |
| $Age^3$ | 0.01 | 0.00 – 0.02 | **0.034** | -26.07 | -41.83 – -10.32 | **0.001** | | | | | | |
| Sex [male]: ref. | | | | | | | | | | | | |

| | | | | | | | | | | | | |
|---|---|---|---|---|---|---|---|---|---|---|---|---|
| Sex [female] | -1.30 | -2.27 – -0.33 | **0.009** | -38.49 | -50.60 – -26.38 | **<0.001** | -8.62 | -16.53 – -0.71 | **0.033** | -0.12 | -0.32 – 0.08 | 0.253 |
| SES [low] | 0.14 | -1.38 – 1.67 | 0.853 | 40.40 | 21.74 – 59.06 | **<0.001** | 11.28 | -0.91 – 23.47 | 0.070 | -0.09 | -0.39 – 0.22 | 0.588 |
| SES [medium]: ref. | | | | | | | | | | | | |
| SES [high] | 2.04 | 0.92 – 3.17 | **<0.001** | -36.48 | -50.65 – -22.30 | **<0.001** | -3.08 | -12.27 – 6.12 | 0.512 | 0.33 | 0.10 – 0.57 | **0.005** |
| BMI [severe underweight] | 1.58 | -1.13 – 4.30 | 0.253 | -4.97 | -38.85 – 28.90 | 0.773 | -7.11 | -29.01 – 14.80 | 0.525 | -0.34 | -0.90 – 0.21 | 0.224 |
| BMI [underweight] | 1.76 | -0.22 – 3.75 | 0.081 | -11.96 | -37.77 – 13.85 | 0.364 | -15.11 | -31.78 – 1.55 | 0.075 | 0.12 | -0.30 – 0.55 | 0.571 |
| BMI [normalweight]: ref. | | | | | | | | | | | | |
| BMI [overweight] | -2.42 | -4.64 – -0.20 | **0.033** | 14.24 | -12.83 – 41.30 | 0.302 | 10.67 | -7.22 – 28.56 | 0.242 | -0.06 | -0.51 – 0.38 | 0.779 |

| | | | | | | | | | | | | |
|---|---|---|---|---|---|---|---|---|---|---|---|---|
| BMI [obese] | 1.20 | -1.76 – 4.16 | 0.428 | 27.42 | -10.34 – 65.17 | 0.155 | -14.76 | -39.46 – 9.94 | 0.241 | -0.25 | -0.86 – 0.37 | 0.437 |
| Covid infection [yes] | -0.29 | -1.41 – 0.83 | 0.610 | -10.44 | -24.26 – 3.37 | 0.138 | -0.05 | -8.82 – 8.71 | 0.991 | -0.18 | -0.40 – 0.04 | 0.118 |
| Covid infection [no]: ref. | | | | | | | | | | | | |
| SDQ | -5.05 | -6.20 – -3.90 | **<0.001** | 41.58 | 26.89 – 56.27 | **<0.001** | -4.68 | -14.20 – 4.84 | 0.335 | -0.35 | -0.59 – -0.11 | **0.005** |
| $SDQ^2$ | 0.48 | -0.86 – 1.81 | 0.484 | -17.37 | -33.89 – -0.86 | **0.039** | -8.45 | -19.22 – 2.32 | 0.124 | -0.15 | -0.42 – 0.13 | 0.293 |
| **Random Effects** | | | | | | | | | | | | |
| $\sigma^2$ | 64.57 | | | 7655.32 | | | 2796.56 | | | 2.72 | | |
| $\tau_{00}$ | 9.40 $_{id}$ | | | 3746.16 $_{id}$ | | | 1780.51 $_{id}$ | | | 0.71 $_{id}$ | | |
| ICC | 0.13 | | | 0.33 | | | 0.39 | | | 0.21 | | |
| N | 806 $_{id}$ | | | 818 $_{id}$ | | | 817 $_{id}$ | | | 818 $_{id}$ | | |
| Observations | 1371 | | | 1626 | | | 1585 | | | 1633 | | |

| | | | | |
|---|---|---|---|---|
| Marginal $R^2$ / Conditional $R^2$ | 0.199 / 0.300 | 0.361 / 0.571 | 0.110 / 0.456 | 0.071 / 0.264 |

Supplement Table 5. Linear mixed model H2 for health related quality of life, recreational screen time, everyday physical activity, and days with >60 minutes moderate-to-vigorous physical activity interaction age (and polynomials) with timepoint, adjusted for sex, socioeconomic status (SES), body mass index (BMI), covid infection, and risk for mental disorder (SDQ) (and polynomials).

| | **Health-related quality of life** | | | **Recreational screen time** | | | **Everyday physical activity** | | | **Days with ≥60 min of moderate to vigorous physical activity** | | |
|---|---|---|---|---|---|---|---|---|---|---|---|---|
| *Predictors* | *Estimates* | *CI* | *p* | *Estimates* | *CI* | *p* | *Estimates* | *CI* | *p* | *Estimates* | *CI* | *p* |
| (Intercept) | 54.05 | 52.73 – 55.38 | **<0.001** | 179.68 | 163.39 – 195.97 | **<0.001** | 116.18 | 105.70 – 126.66 | **<0.001** | 4.36 | 4.09 – 4.62 | **<0.001** |
| Wave [pre-pandemic]: ref. | | | | | | | | | | | | |
| wave [post pandemic] | -5.54 | -6.74 – -4.33 | **<0.001** | 74.25 | 66.13 – 82.37 | **<0.001** | -30.76 | -35.94 – -25.59 | **<0.001** | -0.26 | -0.42 – -0.10 | **0.001** |
| Age | -2.32 | -3.82 – -0.82 | **0.003** | 102.70 | 84.95 – 120.45 | **<0.001** | -5.56 | -6.78 – -4.34 | **<0.001** | -0.12 | -0.15 – -0.08 | **<0.001** |
| Age² | -2.14 | -2.98 – -1.30 | **<0.001** | 8.53 | -0.08 – 17.14 | 0.052 | | | | | | |
| Age³ | 1.61 | -0.11 – 3.32 | 0.067 | -45.04 | -64.06 – -26.03 | **<0.001** | | | | | | |
| Sex[male]: ref. | | | | | | | | | | | | |

| | | | | | | | | | | | | |
|---|---|---|---|---|---|---|---|---|---|---|---|---|
| Sex [female] | -1.35 | -2.32 – -0.38 | **0.006** | -38.46 | -50.57 – -26.35 | **<0.001** | -8.60 | -16.52 – -0.69 | **0.033** | -0.12 | -0.32 – 0.08 | 0.253 |
| SES [low] | 0.13 | -1.39 – 1.65 | 0.869 | 40.24 | 21.58 – 58.90 | **<0.001** | 11.29 | -0.91 – 23.49 | 0.070 | -0.08 | -0.39 – 0.22 | 0.588 |
| SES [medium]: ref. | | | | | | | | | | | | |
| SES [high] | 2.07 | 0.95 – 3.19 | **<0.001** | -36.59 | -50.77 – -22.41 | **<0.001** | -2.89 | -12.09 – 6.32 | 0.538 | 0.33 | 0.10 – 0.57 | **0.005** |
| BMI [severe underweight] | 1.53 | -1.18 – 4.24 | 0.268 | -5.73 | -39.59 – 28.14 | 0.740 | -7.22 | -29.14 – 14.70 | 0.518 | -0.34 | -0.90 – 0.21 | 0.225 |
| BMI [underweight] | 1.80 | -0.18 – 3.78 | 0.075 | -11.55 | -37.36 – 14.25 | 0.380 | -15.10 | -31.78 – 1.58 | 0.076 | 0.12 | -0.30 – 0.55 | 0.571 |
| BMI [normalweight]: ref. | | | | | | | | | | | | |
| BMI [overweight] | -2.47 | -4.68 – -0.25 | **0.029** | 14.20 | -12.87 – 41.26 | 0.304 | 10.70 | -7.20 – 28.60 | 0.241 | -0.06 | -0.51 – 0.38 | 0.779 |

| | | | | | | | | | | | | |
|---|---|---|---|---|---|---|---|---|---|---|---|---|
| BMI [obese] | 1.22 | -1.73 – 4.18 | 0.416 | 27.16 | -10.58 – 64.90 | 0.158 | -15.22 | -39.93 – 9.50 | 0.227 | -0.25 | -0.86 – 0.37 | 0.437 |
| Covid infection [yes] | -0.27 | -1.40 – 0.85 | 0.633 | -10.19 | -24.01 – 3.62 | 0.148 | -0.06 | -8.83 – 8.71 | 0.990 | -0.18 | -0.40 – 0.04 | 0.118 |
| Covid infection [no] | | | | | | | | | | | | |
| SDQ | -5.03 | -6.17 – -3.88 | **<0.001** | 41.90 | 27.21 – 56.58 | **<0.001** | -4.63 | -14.15 – 4.90 | 0.341 | -0.35 | -0.59 – -0.11 | **0.005** |
| SDQ² | 0.52 | -0.81 – 1.86 | 0.442 | -17.15 | -33.67 – -0.63 | **0.042** | -8.50 | -19.27 – 2.27 | 0.122 | -0.15 | -0.42 – 0.13 | 0.293 |
| Wave [pre-pandemic] × age: ref. | | | | | | | | | | | | |
| Wave [post-pandemic] × age | -2.30 | -4.56 – -0.04 | **0.046** | -44.92 | -64.59 – -25.25 | **<0.001** | 3.10 | 1.79 – 4.41 | **<0.001** | 0.02 | -0.02 – 0.06 | 0.433 |
| Wave [post-pandemic] × age² | 1.51 | 0.13 – 2.89 | **0.032** | -38.26 | -47.67 – -28.85 | **<0.001** | | | | | | |

| | | | | | | | | | | |
|---|---|---|---|---|---|---|---|---|---|---|
| Wave [post-pandemic] × age$^3$ | 1.16 | -2.10 – 4.41 | 0.485 | 37.33 | 16.24 – 58.42 | **0.001** | | | | |
| **Random Effects** | | | | | | | | | | |
| $\sigma^2$ | 63.59 | | | 6935.56 | | | 2720.49 | | | 2.73 |
| $\tau_{00}$ | 9.71 $_{id}$ | | | 4108.03 $_{id}$ | | | 1824.25 $_{id}$ | | | 0.71 $_{id}$ |
| ICC | 0.13 | | | 0.37 | | | 0.40 | | | 0.21 |
| N | 806 $_{id}$ | | | 818 $_{id}$ | | | 817 $_{id}$ | | | 818 $_{id}$ |
| Observations | 1371 | | | 1626 | | | 1585 | | | 1633 |
| Marginal $R^2$ / Conditional $R^2$ | 0.207 / 0.312 | | | 0.382 / 0.612 | | | 0.117 / 0.472 | | | 0.072 / 0.264 |

Supplement Table 6. Linear mixed model health related quality of life (HrQoL), recreational screen time (ST), everyday physical activity (Everyday PA), and days with >60 minutes moderate-to-vigorous physical activity (WHO PA days) for the measurements pre pandemic, lockdown 1 (LD1), lockdown 2 (LD2), and post pandemic adjusted for age centered (and polynomials), sex, socioeconomic status (SES), body mass index (BMI), covid infection, and risk for mental disorder (SDQ) (and polynomials).

| | **Health-related quality of life** | | | **Recreational screen time** | | | **Everyday physical activity** | | | **Days with ≥60 min of moderate to vigorous physical activity** | | |
|---|---|---|---|---|---|---|---|---|---|---|---|---|
| *Predictors* | *Estimates* | *CI* | *p* | *Estimates* | *CI* | *p* | *Estimates* | *CI* | *p* | *Estimates* | *CI* | *p* |
| (Intercept) | 49.31 | 47.38 – 51.24 | **<0.001** | 274.78 | 244.41 – 305.15 | **<0.001** | 108.33 | 92.34 – 124.31 | **<0.001** | 4.52 | 4.13 – 4.92 | **<0.001** |
| Wave [pre-pandemic to LD1] | -10.36 | -11.14 – -9.57 | **<0.001** | 61.13 | 51.99 – 70.26 | **<0.001** | 37.07 | 30.54 – 43.61 | **<0.001** | 0.41 | 0.24 – 0.58 | **<0.001** |
| Wave [LD1 to LD2] | -0.12 | -0.89 – 0.65 | 0.763 | 39.85 | 30.74 – 48.97 | **<0.001** | -82.54 | -89.12 – -75.96 | **<0.001** | -0.80 | -0.96 – -0.63 | **<0.001** |
| Wave [LD2 to post-pandemic] | 5.08 | 4.32 – 5.85 | **<0.001** | -26.02 | -34.62 – -17.43 | **<0.001** | 14.42 | 8.12 – 20.72 | **<0.001** | 0.12 | -0.04 – 0.28 | 0.136 |
| Age | -0.55 | -0.80 – -0.31 | **<0.001** | 23.88 | 19.98 – 27.78 | **<0.001** | -4.28 | -5.26 – -3.30 | **<0.001** | -0.13 | -0.15 – -0.10 | **<0.001** |
| $Age^2$ | -0.07 | -0.10 – -0.03 | **<0.001** | -0.66 | -1.18 – -0.15 | **0.012** | | | | | | |
| $Age^3$ | 0.01 | 0.00 – 0.02 | **0.006** | -0.26 | -0.39 – -0.13 | **<0.001** | | | | | | |
| Sex[male]: ref. | | | | | | | | | | | | |

| | | | | | | | | | | | | |
|---|---|---|---|---|---|---|---|---|---|---|---|---|
| Sex [female] | -1.09 | -1.90 – -0.28 | **0.008** | -34.77 | -47.49 – -22.06 | **<0.001** | -8.19 | -15.67 – -0.72 | **0.032** | -0.06 | -0.25 – 0.12 | 0.509 |
| SES [low] | -0.23 | -1.49 – 1.03 | 0.718 | 37.80 | 18.16 – 57.43 | **<0.001** | 11.94 | 0.42 – 23.47 | **0.042** | -0.02 | -0.31 – 0.27 | 0.888 |
| SES [medium]: ref. | | | | | | | | | | | | |
| SES [high] | 1.91 | 0.97 – 2.86 | **<0.001** | -36.50 | -51.38 – -21.61 | **<0.001** | -3.07 | -11.74 – 5.61 | 0.488 | 0.37 | 0.15 – 0.58 | **0.001** |
| BMI [severe underweight] | 1.90 | -0.35 – 4.15 | 0.097 | -11.72 | -47.16 – 23.71 | 0.517 | -0.66 | -21.44 – 20.11 | 0.950 | -0.43 | -0.95 – 0.09 | 0.102 |
| BMI [underweight] | 1.08 | -0.60 – 2.77 | 0.208 | -5.24 | -32.20 – 21.72 | 0.703 | -13.30 | -29.02 – 2.43 | 0.097 | -0.08 | -0.48 – 0.31 | 0.677 |
| BMI [normalweight]: ref. | | | | | | | | | | | | |
| BMI [overweight] | -0.75 | -2.56 – 1.06 | 0.417 | 14.81 | -13.58 – 43.21 | 0.306 | 13.53 | -3.11 – 30.18 | 0.111 | -0.03 | -0.44 – 0.39 | 0.892 |
| BMI [obese] | 0.77 | -1.72 – 3.26 | 0.543 | 35.61 | -3.93 – 75.15 | 0.077 | -18.03 | -41.27 – 5.20 | 0.128 | -0.32 | -0.90 – 0.26 | 0.276 |
| Covid infection [yes] | -0.35 | -1.27 – 0.57 | 0.459 | -18.18 | -32.68 – -3.67 | **0.014** | -1.87 | -10.14 – 6.41 | 0.658 | -0.23 | -0.44 – -0.03 | **0.026** |

| | | | | | | | | | | | | |
|---|---|---|---|---|---|---|---|---|---|---|---|---|
| Covid infection [no] | | | | | | | | | | | | |
| SDQ | -4.14 | -5.10 – -3.17 | **<0.001** | 43.77 | 28.40 – 59.14 | **<0.001** | -5.92 | -14.94 – 3.09 | 0.198 | -0.31 | -0.54 – -0.09 | **0.006** |
| SDQ² | 0.31 | -0.80 – 1.41 | 0.589 | -13.27 | -30.60 – 4.06 | 0.133 | -8.05 | -18.20 – 2.10 | 0.120 | -0.03 | -0.28 – 0.23 | 0.832 |
| **Random Effects** | | | | | | | | | | | | |
| $\sigma^2$ | 52.32 | | | 7839.51 | | | 4010.06 | | | 2.64 | | |
| $\tau_{00}$ | 18.11 id | | | 6320.16 id | | | 1808.72 id | | | 1.11 id | | |
| ICC | 0.26 | | | 0.45 | | | 0.31 | | | 0.30 | | |
| N | 818 id | | | 818 id | | | 818 id | | | 818 id | | |
| Observations | 2782 | | | 3122 | | | 3050 | | | 3131 | | |
| Marginal $R^2$ / Conditional $R^2$ | 0.269 / 0.457 | | | 0.350 / 0.640 | | | 0.184 / 0.438 | | | 0.098 / 0.365 | | |

Supplement Table 7. Linear mixed model health related quality of life (HrQoL), recreational screen time (ST), everyday physical activity (Everyday PA), and days with >60 minutes moderate-to-vigorous physical activity (WHO PA days) for the measurements pre pandemic, lockdown 1 (LD1), lockdown 2 (LD2), and post pandemic and their interaction with age, adjusted for age centered (and polynomials), sex, socioeconomic status (SES), body mass index (BMI), covid infection, and risk for mental disorder (SDQ) (and polynomials).

| | **Health-related quality of life** | | | **Recreational screen time** | | | **Everyday physical activity** | | | **Days with ≥60 min of moderate to vigorous physical activity** | | |
|---|---|---|---|---|---|---|---|---|---|---|---|---|
| *Predictors* | *Estimates* | *CI* | *p* | *Estimates* | *CI* | *p* | *Estimates* | *CI* | *p* | *Estimates* | *CI* | *p* |
| (Intercept) | 49.13 | 47.20 – 51.07 | **<0.001** | 273.53 | 243.16 – 303.91 | **<0.001** | 108.52 | 92.58 – 124.45 | **<0.001** | 4.53 | 4.13 – 4.93 | **<0.001** |
| Wave [pre-pandemic to LD1] | -11.97 | -13.12 – -10.82 | **<0.001** | 66.63 | 53.01 – 80.25 | **<0.001** | 36.28 | 29.92 – 42.64 | **<0.001** | 0.40 | 0.23 – 0.57 | **<0.001** |
| Wave [LD1 to LD2] | 0.60 | -0.55 – 1.74 | 0.307 | 59.44 | 45.86 – 73.03 | **<0.001** | -81.19 | -87.58 – -74.79 | **<0.001** | -0.79 | -0.95 – -0.62 | **<0.001** |
| Wave [LD2 to post-pandemic] | 3.99 | 2.85 – 5.14 | **<0.001** | -15.21 | -27.94 – -2.47 | **0.019** | 14.23 | 8.11 – 20.35 | **<0.001** | 0.12 | -0.03 – 0.28 | 0.122 |
| Age | -0.58 | -0.83 – -0.33 | **<0.001** | 24.08 | 20.18 – 27.98 | **<0.001** | -4.47 | -5.45 – -3.50 | **<0.001** | -0.13 | -0.16 – -0.11 | **<0.001** |
| $Age^2$ | -0.07 | -0.11 – -0.03 | **<0.001** | -0.61 | -1.13 – -0.10 | **0.020** | | | | | | |
| $Age^3$ | 0.01 | 0.00 – 0.02 | **0.004** | -0.26 | -0.39 – -0.14 | **<0.001** | | | | | | |
| Sex [male]: ref. | | | | | | | | | | | | |

| | | | | | | | | | | | | |
|---|---|---|---|---|---|---|---|---|---|---|---|---|
| Sex [female] | -1.11 | -1.92 – -0.31 | **0.007** | -34.91 | -47.63 – -22.19 | **<0.001** | -8.13 | -15.58 – -0.68 | **0.032** | -0.06 | -0.25 – 0.12 | 0.511 |
| SES [low] | -0.19 | -1.45 – 1.07 | 0.767 | 38.09 | 18.45 – 57.72 | **<0.001** | 11.46 | -0.03 – 22.94 | 0.051 | -0.02 | -0.31 – 0.26 | 0.867 |
| SES [medium]: ref. | | | | | | | | | | | | |
| SES [high] | 1.97 | 1.03 – 2.91 | **<0.001** | -36.25 | -51.14 – -21.36 | **<0.001** | -3.26 | -11.91 – 5.39 | 0.460 | 0.36 | 0.15 – 0.58 | **0.001** |
| BMI [severe underweight] | 1.88 | -0.37 – 4.13 | 0.101 | -12.06 | -47.50 – 23.38 | 0.505 | -0.91 | -21.62 – 19.80 | 0.931 | -0.43 | -0.95 – 0.08 | 0.101 |
| BMI [underweight] | 1.11 | -0.58 – 2.80 | 0.198 | -4.85 | -31.82 – 22.11 | 0.724 | -13.42 | -29.10 – 2.26 | 0.093 | -0.08 | -0.48 – 0.31 | 0.674 |
| BMI [normalweight]: ref. | | | | | | | | | | | | |
| BMI [overweight] | -0.79 | -2.60 – 1.02 | 0.392 | 14.96 | -13.44 – 43.36 | 0.302 | 13.82 | -2.77 – 30.42 | 0.102 | -0.03 | -0.44 – 0.39 | 0.898 |
| BMI [obese] | 0.85 | -1.64 – 3.34 | 0.502 | 35.52 | -4.01 – 75.06 | 0.078 | -18.31 | -41.47 – 4.85 | 0.121 | -0.32 | -0.90 – 0.26 | 0.275 |
| Covid infection [yes] | -0.28 | -1.20 – 0.65 | 0.555 | -17.92 | -32.43 – -3.42 | **0.015** | -1.98 | -10.23 – 6.27 | 0.638 | -0.24 | -0.44 – -0.03 | **0.026** |

| | | | | | | | | | | | | |
|---|---|---|---|---|---|---|---|---|---|---|---|---|
| Covid infection [no]: ref. | | | | | | | | | | | | |
| SDQ | -4.13 | -5.10 – -3.16 | **<0.001** | 43.76 | 28.39 – 59.12 | **<0.001** | -5.73 | -14.72 – 3.25 | 0.211 | -0.31 | -0.54 – -0.09 | **0.006** |
| SDQ² | 0.35 | -0.76 – 1.45 | 0.541 | -13.00 | -30.33 – 4.33 | 0.141 | -7.98 | -18.11 – 2.14 | 0.122 | -0.03 | -0.28 – 0.23 | 0.834 |
| Wave [pre-pandemic to LD1] x age | 0.50 | 0.04 – 0.97 | **0.033** | 1.08 | -4.37 – 6.53 | 0.698 | -3.87 | -5.48 – -2.26 | **<0.001** | -0.05 | -0.09 – -0.01 | **0.017** |
| Wave [LD1 to LD2] x age | -0.36 | -0.82 – 0.10 | 0.128 | 1.83 | -3.61 – 7.26 | 0.510 | 9.29 | 7.67 – 10.90 | **<0.001** | 0.04 | -0.00 – 0.08 | 0.071 |
| Wave [LD2 to post-pandemic] x age | -0.81 | -1.31 – -0.30 | **0.002** | -14.44 | -19.55 – -9.33 | **<0.001** | -2.39 | -3.94 – -0.83 | **0.003** | 0.03 | -0.01 – 0.07 | 0.167 |
| Wave [pre-pandemic to LD1] × age² | 0.12 | 0.05 – 0.18 | **0.001** | -0.39 | -1.10 – 0.32 | 0.281 | | | | | | |
| Wave [LD1 to LD2] x age² | -0.05 | -0.11 – 0.01 | 0.128 | -1.22 | -1.92 – -0.51 | **0.001** | | | | | | |
| Wave [LD2 to post-pandemic] x age² | 0.04 | -0.04 – 0.13 | 0.297 | -1.02 | -1.68 – -0.35 | **0.003** | | | | | | |
| Wave [pre-pandemic to LD1] × age³ | -0.01 | -0.02 – 0.01 | 0.525 | 0.06 | -0.12 – 0.24 | 0.512 | | | | | | |

| | | | | | | | | | | |
|---|---|---|---|---|---|---|---|---|---|---|
| Wave [LD1 to LD2] x age$^3$ | 0.00 | -0.01 – 0.02 | 0.703 | -0.04 | -0.22 – 0.14 | 0.654 | | | | |
| Wave [LD2 to post-pandemic] x age$^3$ | 0.02 | -0.01 – 0.04 | 0.178 | 0.28 | 0.11 – 0.45 | **0.001** | | | | |
| **Random Effects** | | | | | | | | | | |
| $\sigma^2$ | 50.59 | | | 7419.70 | | | 3783.13 | | | 2.63 |
| $\tau_{00}$ | 18.61 $_{id}$ | | | 6431.47 $_{id}$ | | | 1851.81 $_{id}$ | | | 1.11 $_{id}$ |
| ICC | 0.27 | | | 0.46 | | | 0.33 | | | 0.30 |
| N | 818 $_{id}$ | | | 818 $_{id}$ | | | 818 $_{id}$ | | | 818 $_{id}$ |
| Observations | 2782 | | | 3122 | | | 3050 | | | 3131 |
| Marginal $R^2$ / Conditional $R^2$ | 0.283 / 0.476 | | | 0.366 / 0.660 | | | 0.210 / 0.469 | | | 0.100 / 0.367 |
